# Sensitivity of Rapid Antigen Tests Against SARS-CoV-2 Omicron and Delta Variants

**DOI:** 10.1101/2023.02.09.23285583

**Authors:** Anuradha Rao, Adrianna Westbrook, Leda Bassit, Richard Parsons, Eric Fitts, Morgan Greenleaf, Kaleb McLendon, Julie A. Sullivan, William O’Sick, Tyler Baugh, Heather B. Bowers, Filipp Frank, Ethan Wang, Mimi Le, Jennifer Frediani, Pavitra Roychoudhury, Alexander L. Greninger, Robert Jerris, Nira R. Pollock, Eric A. Ortlund, John D. Roback, Wilbur A. Lam, Anne Piantadosi

**Author notes:** Co-corresponding authors **Co-Corresponding authors:** Wilbur A. Lam, 2015, Uppergate Drive, Atlanta, GA 30322, Anne Piantadosi, 101 Woodruff Circle 7207A Atlanta, GA 30322. Contributed equally to this manuscript (Anuradha Rao, Adrianna Westbrook).

## Abstract

Rapid Antigen Tests (RAT) have become an invaluable tool for combating the COVID-19 pandemic. However, concerns have been raised regarding the ability of existing RATs to effectively detect emerging SARS-CoV-2 variants. We compared the performance of eight commercially available, emergency use authorized RATs against the Delta and Omicron SARS-CoV-2 variants using individual patient and serially diluted pooled clinical samples. The RATs exhibited lower sensitivity for Omicron samples when using PCR Cycle threshold (C_T_) value (a proxy for RNA concentration) as the comparator. Interestingly, however, they exhibited similar sensitivity for Omicron and Delta samples when using quantitative antigen concentration as the comparator. We further found that the Omicron samples had lower ratios of antigen to RNA, which offers a potential explanation for the apparent lower sensitivity of RATs for that variant when using C_T_ value as a reference. Our findings underscore the complexity in assessing RAT performance against emerging variants and highlight the need for ongoing evaluation in the face of changing population immunity and virus evolution.

## INTRODUCTION

As the SARS-CoV-2 pandemic progresses, rapid antigen tests (RATs) have become a key component of home testing, community screening, and clinical diagnostics owing to their ease of use, low cost, and speed. In the United States, there are currently 19 over-the-counter antigen tests and 23 point-of-care antigen tests available under Emergency Use Authorization (EUA), and hundreds of millions of antigen tests are used every month (1). Concurrently with these important advances in the availability, variety, and widespread use of RATs, SARS-CoV-2 continues to evolve, raising concern that new variants may harbor genetic and antigenic changes affecting test performance. The Omicron variant, which was first reported in November 2021 and quickly replaced Delta as the predominant variant in the U.S., differs from Delta by 7 amino acid changes and a 2-amino acid deletion in the nucleocapsid (N) protein, the target of most RATs. Prior studies have demonstrated conflicting results, with some showing decreased performance of RATs for Omicron, but others showing comparable performance (2, 3). We compared the performance characteristics of eight RATs in detecting Delta and Omicron variants, using both individual clinical samples and standardized pools of clinical samples, and used orthogonal protein detection, RNA detection, and infectivity measurements to understand variant-specific differences in RAT results.

## RESULTS

### Rapid antigen test sensitivities for Delta and Omicron using serially diluted, pooled clinical samples are similar when using antigen concentration as the comparator, but not when using RNA measured by cycle threshold (C_T_) value as the comparator

We evaluated the sensitivity of eight commercially available RATs for Omicron and Delta using a standardized set of pooled remnant clinical samples (RCS pools) that were serially diluted and quantified for SARS-CoV-2 RNA (measured by C_T_ value from CDC N2 PCR assay; described in supplementary methods) and nucleocapsid antigen concentration (measured by Quanterix Simoa Assay; described in supplementary methods). When RAT limit of detection (LoD) was measured using antigen concentration as comparator, only the BinaxNow assay was less sensitive in detecting Omicron than Delta, with a three-fold higher LoD (**Figure 1A, Figure S1**). The other tests performed similarly against Delta and Omicron pools, with a twofold or less difference in LoD (**Figure 1A, Figure S1, Figure S2**). However, when LoD was measured using C_T_ value as a comparator, 5 of the 8 RATs were less sensitive in detecting Omicron than Delta (LoD C_T_ difference ranging from 2.5-3.2 lower for Omicron, corresponding to a nearly tenfold higher RNA concentration) (**Figure 1B, Figure S1, Figure S2**).

**Fig. 1.**
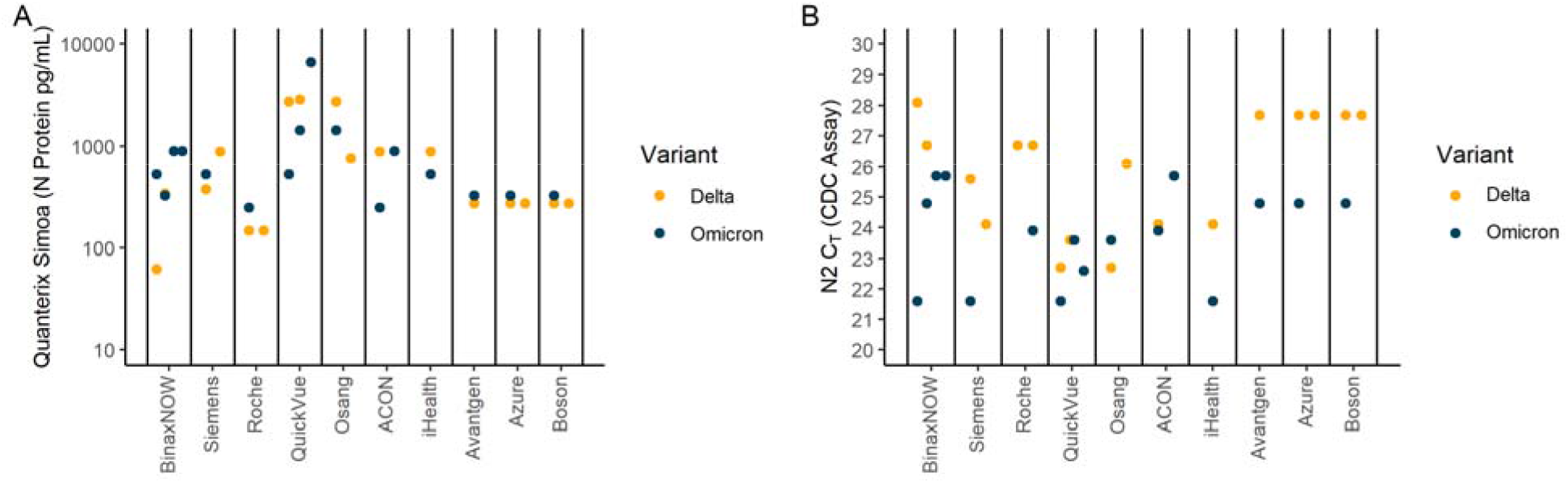
Results of testing 8 commercially available rapid antigen tests against remnant clinical sample pools. Each of eight commercially available rapid antigen tests (RATs) was tested using sequence-confirmed, serially diluted and quantified (N2 C_T_ CDC Assay and Quanterix Simoa SARS-CoV-2 N Protein Antigen Test) pools generated from remnant clinical samples (RCS) of the same variant. Panels A and B show each RAT’s limit of detection, which is the lowest antigen concentration **(A)** or highest C_T_ value **(B)** that was detected in five out of five replicates. When testing was repeated with independent RCS pools, or independent lots of tests, they are presented as separate data points.

Thus, using RCS pools, we observed concordant sensitivity for Omicron and Delta across most commercially available RATs when measured against antigen concentration, but lower sensitivity for Omicron than Delta for over half of the RATs when measured against C_T_ value, suggesting different relationships between antigen concentration and RNA concentration for Delta versus Omicron.

### Rapid antigen test sensitivity for Delta and Omicron using individual clinical samples varies between assays and depends on choice of comparator

We next evaluated the sensitivity of two common RATs using a subset of anterior nares specimens from a large study in which 171 fresh remnant clinical samples (RCS) were collected from individuals infected with Omicron and 163 banked RCS had been collected from individuals infected with Delta (**Table 1, Supplementary Data File**). Many of the participants were unvaccinated (40.9% and 48.5% of those infected with Delta and Omicron, respectively), and individuals infected with Omicron had shorter times since last vaccine dose (**Table 1**). Per study design, most participants were symptomatic (**Table 1**). However, individuals infected with Omicron had shorter durations of symptoms prior to testing: nearly 80% were tested within 3 days of symptom onset and the remaining 20% within 7 days. By contrast, only about a quarter of patients with Delta were tested within 3 days of symptom onset, about half between 3 and 7 days, and about a quarter after 7 days.

**Table 1.** Demographics of patients with Delta (N=163) and Omicron (N=171).

|  | **Delta, n (%)** | **Omicron, n (%)** |
| --- | --- | --- |
| **Sex** |  |  |
| Female | 98 (60.9%) | 91 (53.2%) |
| Male | 63 (39.1%) | 80 (46.8%) |
| **Race** |  |  |
| White | 79 (47.9%) | 80 (44.9%) |
| Black/African American | 73 (44.2%) | 75 (42.1%) |
| Asian | 3 (1.8%) | 8 (4.5%) |
| Other | 10 (6.1%) | 11 (6.2%) |
| Refuse to Answer | 0 (0%) | 4 (2.3%) |
| **Ethnicity** |  |  |
| Hispanic | 12 (7.4%) | 26 (15.2%) |
| Non-Hispanic | 149 (92.6%) | 144 (84.2%) |
| Refuse to Answer | 0 (0%) | 1 (0.6%) |
| **Vaccine Status** |  |  |
| Unvaccinated/Not Fully Vaccinated | 65 (40.9%) | 83 (48.5%) |
| Fully Vaccinated/Boosted | 94 (59.1%) | 88 (51.5%) |
| **Days Since Last Vaccine** |  |  |
| Within The Last 90 Days | 17 (17.7%) | 41 (43.1%) |
| Between 91 and 180 Days | 22 (22.9%) | 16 (16.8%) |
| Between 181 and 270 Days | 52 (54.2%) | 28 (29.5%) |
| More than 270 Days | 5 (5.2%) | 10 (10.5%) |
| Symptom Status |  |  |
| Asymptomatic | 5 (3.1%) | 1 (0.6%) |
| Symptomatic | 156 (96.9%) | 170 (99.4%) |
| Symptom Duration |  |  |
| Symptoms for at most 3 days | 36 (23.1%) | 132 (77.7%) |
| Symptoms for between 4 and 7 days | 80 (51.3%) | 38 (22.3%) |
| Symptoms for more than 7 days | 40 (25.6%) | 0 (0%) |

From this study population, 75 Delta and 84 Omicron samples with C_T_ less than 30 (Cepheid Xpert assay; described in supplementary methods) were randomly selected for testing with the Abbott BinaxNOW™ COVID-19 Antigen Test and Quidel QuickVue SARS Antigen Test RATs. Across all samples, sensitivity was similar between Delta and Omicron for Quickvue. Sensitivity appeared lower for Omicron than Delta samples for BinaxNow, although this difference was not statistically significant (**Table 2**). As expected, tests were more sensitive in samples with higher concentrations of viral antigen and lower C_T_ values (**Figure 2 A-D**).

**Table 2:** Sensitivity of BinaxNOW and QuickVue RATs using individual RCS. Samples were stratified by antigen concentration (top panel) or C_T_ value (bottom panel), and the sensitivity of detection with corresponding 95% confidence intervals for Delta and Omicron was compared within each stratum using chi-square or Fisher’s exact test. The supplementary data file contains results of BinaxNOW and QuickVue for each observation.

|  | BinaxNow |  |  | QuickVue |  |  |
| --- | --- | --- | --- | --- | --- | --- |
|  | Delta | Omicron | P-value | Delta | Omicron | P-value |
| Overall | 57.3<br>(56.2, 58.4) | 45.7<br>(44.7, 46.8) | 0.18 | 60<br>(58.9, 61.1) | 58.5<br>(57.5, 59.5) | 0.97 |
| **Antigen Concentration (pg/mL)** |  |  |  |  |  |  |
| 10 | 0 (0, 0) | 0 (0, 0) | -- | 0 (0, 0) | 0 (0, 0) | -- |
| 100 | 0 (0, 0) | 0 (0, 0) | -- | 0<br>(0, 0) | 7.4<br>(0.0, 17.3) | 0.52 |
| 1000 | 29.3<br>(15.3, 43.2) | 22.0<br>(11.46, 32.61) | 0.56 | 31.7<br>(17.5, 100) | 37.9<br>(25.4, 50.4) | 0.67 |
| 10000 | 51.5<br>(39.5, 63.6) | 35.4<br>(24.9, 46.0) | 0.08 | 54.6<br>(42.5, 66.6) | 51.3<br>(40.2, 62.4) | 0.74 |
| 100000 | 56.8<br>(45.5, 68.0) | 44.6<br>(34.4, 54.7) | 0.16 | 59.5<br>(48.3, 70.6) | 58.2<br>(48.11, 68.4) | 1 |
| **C<sub>T</sub> Value** |  |  |  |  |  |  |
| ≤20 | 100<br>(100, 100) | 91.7<br>(90.6, 100) | 0.89 | 100<br>(100, 100) | 95.8<br>(95.0, 100) | 1 |
| ≤22 | 95 | 72.6 | 0.08 | 95 | 92.2 | 1 |
|  | (94.0, 100) | (71.3, 73.8) |  | (94.0, 100) | (91.4, 100) |  |
| ≤24 | 93.3 | 58.0 | 0.001 | 93.3 | 76.8 | 0.09 |
|  | (92.4, 100) | (56.8, 59.1) |  | (92.4, 100) | (75.8, 77.8) |  |
| ≤26 | 81.4 | 54.4 | 0.01 | 88.4 | 69.6 | 0.03 |
|  | (80.2, 82.6) | (53.3, 55.5) |  | (87.4, 100) | (68.6, 70.6) |  |
| ≤28 | 70 | 48.3 | 0.01 | 73.3 | 61.8 | 0.16 |
|  | (68.8, 71.2) | (47.3, 49.4) |  | (72.2, 74.4) | (60.8, 62.8) |  |

**Fig. 2.**
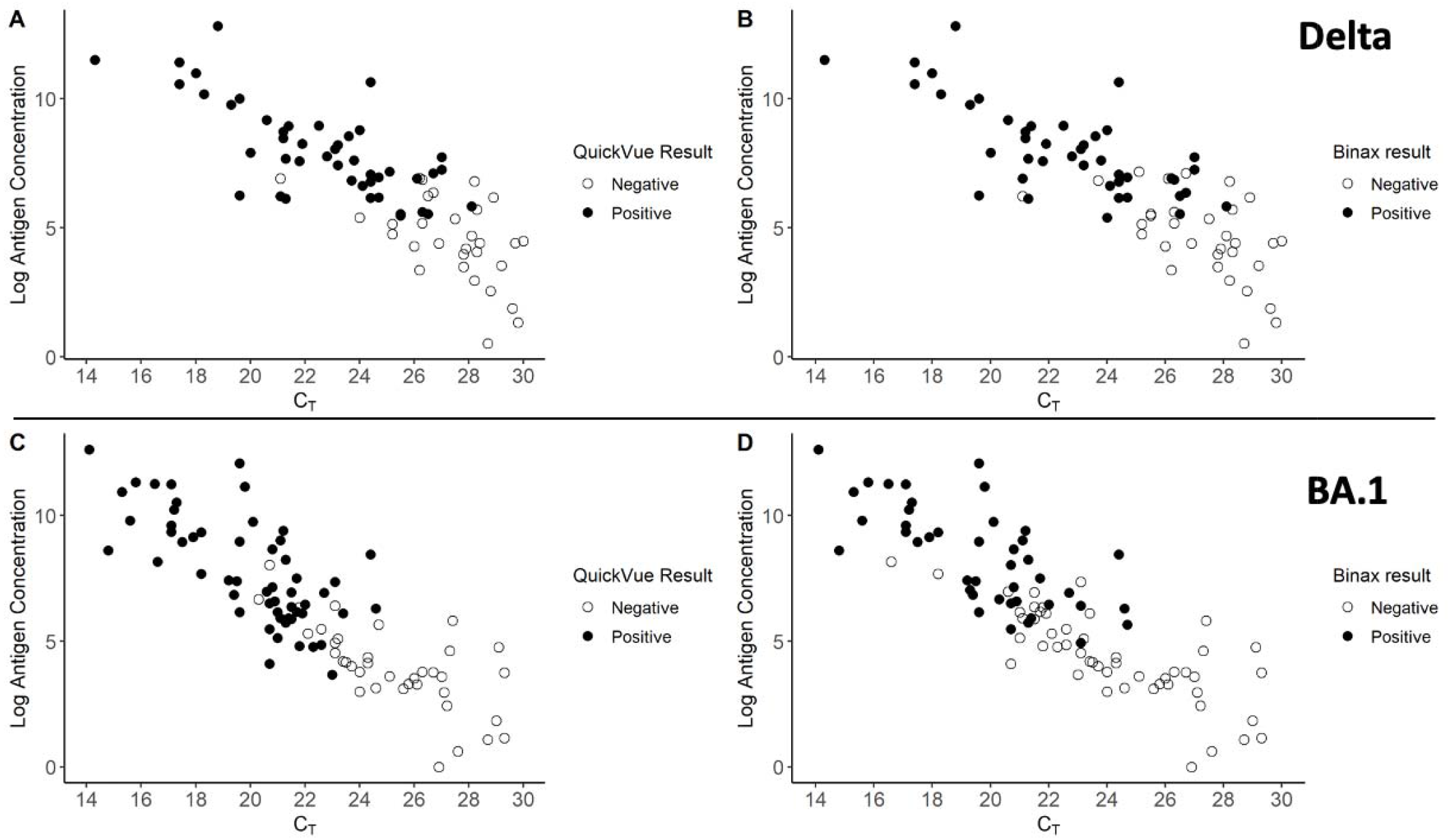
Results of testing 2 commercially available rapid antigen tests against individual remnant clinical samples for Delta (A and B) and Omicron (C and D). Sequence-verified residual mid turbinate samples from 75 individuals with Delta infection and 84 individuals with BA.1 infection underwent RT-PCR testing using the Xpert Xpress CoV-2/Flu/RSV plus & Xpert Xpress SARS-CoV-2 assays (Cepheid), protein quantification using the Simoa SARS-CoV-2 N Protein Antigen assay (Quanterix), and rapid antigen testing (QuickVue or Binax) according to the manufacturer’s instructions. Y-axes reflect natural log(n+1) transformed antigen concentration. Abbreviations: Cycle threshold (C_T_)

When samples were stratified by antigen concentration (**Table 2, top panel**), QuickVue had similar sensitivity for Delta and Omicron across all strata. BinaxNow appeared somewhat less sensitive for Omicron than Delta, but this difference was not statistically significant. For both assays and both variants, sensitivity increased as antigen concentration increased, as expected. When samples were stratified by C_T_ value, sensitivity decreased as C_T_ value increased, as expected (**Table 2, bottom panel**). Both assays showed lower sensitivity for Omicron than Delta for C_T_ thresholds 24 and higher; this result was more pronounced, and only statistically significant, for BinaxNOW.

Thus, results from both individual RCS and RCS pools show that the QuickVue assay has similar sensitivity in detecting Delta and Omicron when antigen concentration is used as a comparator; it has a somewhat (but not statistically significant) reduced sensitivity for Omicron when using C_T_ value as a comparator. The BinaxNOW assay has a somewhat (but not statistically significant) reduced sensitivity for Omicron when antigen concentration is used as a comparator; it has a more pronounced (and statistically significant) reduction in sensitivity for Omicron when C_T_ value is used as a comparator. Results for both individual RCS and RCS pools showed a discrepancy in RAT sensitivity when using antigen concentration versus C_T_ value as the comparator.

To identify potential mutations that might affect test performance, we analyzed SARS-CoV-2 genome sequences. The Delta samples represented a range of sublineages (**Supplementary Data File**). In the N protein, aside from the four lineage-defining mutations (D63G, R203M, G215C, and D377Y), no mutation was present in more than 3 samples. The Omicron samples all belonged to lineage BA.1 or BA.1.1. In addition to lineage-defining mutations (P13L, R203K, G204R, and DEL31-33), 25 of the 152 samples had D343G and 4 had P67S. The Omicron lineages that have emerged since the time of this study, BA.2, BA.4, and BA.5, contain the additional mutation S413R, which was not present in any of these samples. Overall, sequence analysis confirmed that these clinical samples were representative of Delta and Omicron variants and suggested that, if N protein mutations affect test sensitivity, they are likely to be lineage-defining mutations.

### Omicron samples have lower antigen-per-RNA than Delta samples

We formally compared the relationship between antigen concentration and C_T_ in 163 Delta and 169 Omicron individual RCS, including the samples tested by RAT. As expected, antigen concentration and C_T_ were highly correlated, both across all samples and for each variant individually (**Figure 3**), including in sensitivity analysis (**Figure S3**). Notably, regression analysis indicated a significant association between C_T_ value and variant (**Table 3**). Specifically, Omicron samples had a 6.8 (standard error [SE]=0.55) cycle lower C_T_ than Delta samples, for a given antigen concentration (p-value<0.001), indicating a greater amount of RNA-per-antigen (a lower amount of antigen-per-RNA) than Delta samples. In the full regression model that also included vaccine status and presence of symptoms, Omicron samples had a 6.5 (SE=0.57, p-value<0.001) cycle lower C_T_ than Delta samples, for a given antigen concentration. Unsurprisingly, C_T_ value was significantly associated with antigen concentration, with a decrease of 2.3 (SE=0.08, p-value<0.001) cycles and 2.2 (SE=0.08, p-value<0.001) cycles per natural log change in antigen concentration in the base model and full model, respectively. C_T_ value was also significantly associated with the presence of symptoms; it was 8.0 (SE=2.19, p-value<0.001) cycles lower for symptomatic individuals than asymptomatic individuals. We did not observe a significant association between C_T_ value and vaccine status (Beta=0.75, SE=1.34, p-value=0.60). In sensitivity analysis, these relationships remained similar and statistically significant, but with lower magnitude of effect (**Table S1**).

**Table 3.**
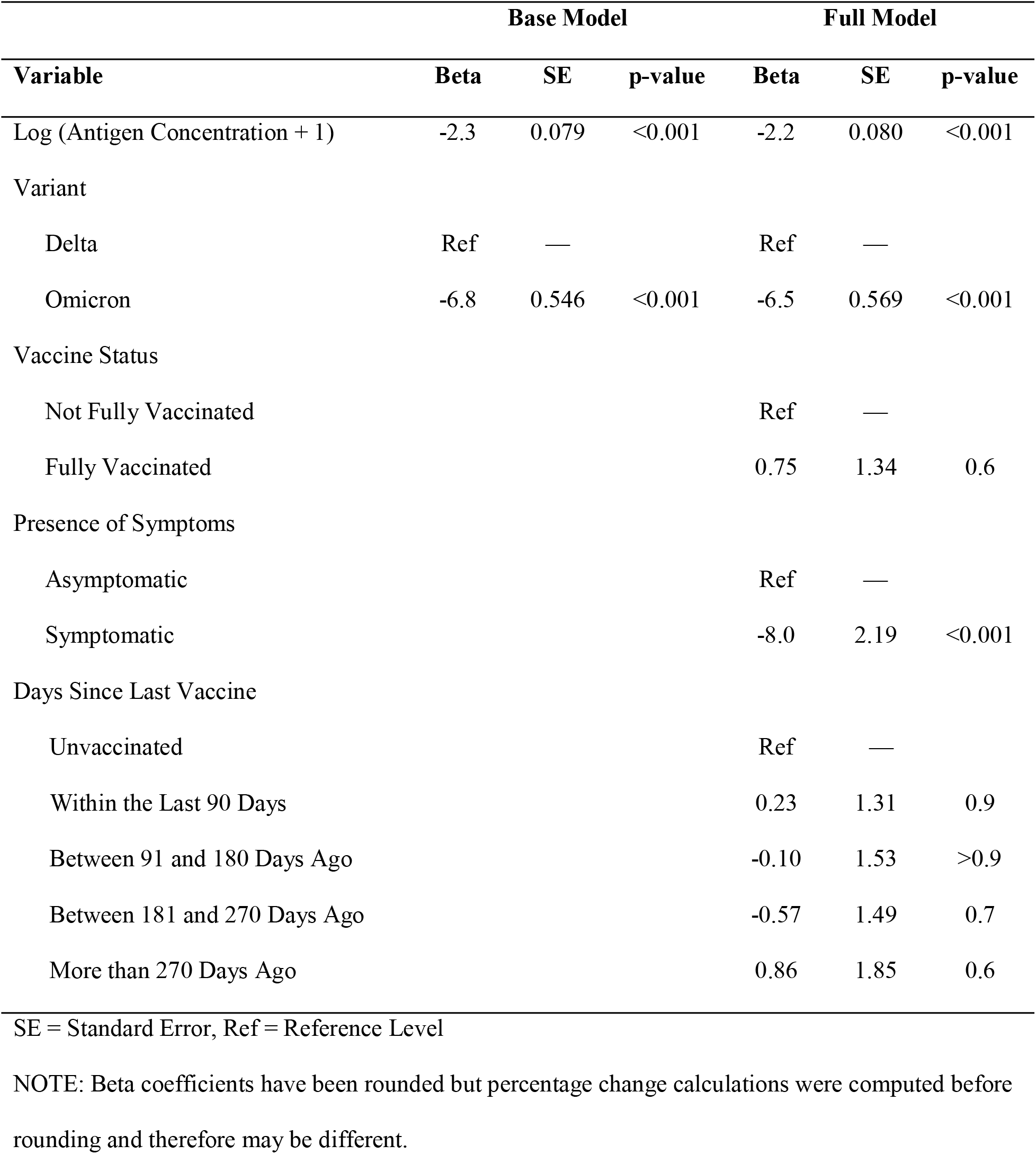
**Association between C**_**T**_ **value and natural log(n+1) transformed antigen concentration (pg/ml), variant, vaccine status, and presence of symptoms** for 151 Delta and 168 Omicron samples, excluding samples with missing information on days since last vaccine or symptom duration.

| Variable | Base Model |  |  | Full Model |  |  |
| --- | --- | --- | --- | --- | --- | --- |
|  | Beta | SE | p-value | Beta | SE | p-value |
| Log (Antigen Concentration + 1) | -2.3 | 0.079 | <0.001 | -2.2 | 0.080 | <0.001 |
| Variant |  |  |  |  |  |  |
| Delta | Ref | — |  | Ref | — |  |
| Omicron | -6.8 | 0.546 | <0.001 | -6.5 | 0.569 | <0.001 |
| Vaccine Status |  |  |  |  |  |  |
| Not Fully Vaccinated |  |  |  | Ref | — |  |
| Fully Vaccinated |  |  |  | 0.75 | 1.34 | 0.6 |
| Presence of Symptoms |  |  |  |  |  |  |
| Asymptomatic |  |  |  | Ref | — |  |
| Symptomatic |  |  |  | -8.0 | 2.19 | <0.001 |
| Days Since Last Vaccine |  |  |  |  |  |  |
| Unvaccinated |  |  |  | Ref | — |  |
| Within the Last 90 Days |  |  |  | 0.23 | 1.31 | 0.9 |
| Between 91 and 180 Days Ago |  |  |  | -0.10 | 1.53 | >0.9 |
| Between 181 and 270 Days Ago |  |  |  | -0.57 | 1.49 | 0.7 |
| More than 270 Days Ago |  |  |  | 0.86 | 1.85 | 0.6 |

SE = Standard Error, Ref = Reference Level
NOTE: Beta coefficients have been rounded but percentage change calculations were computed before rounding and therefore may be different.

**Fig. 3.**
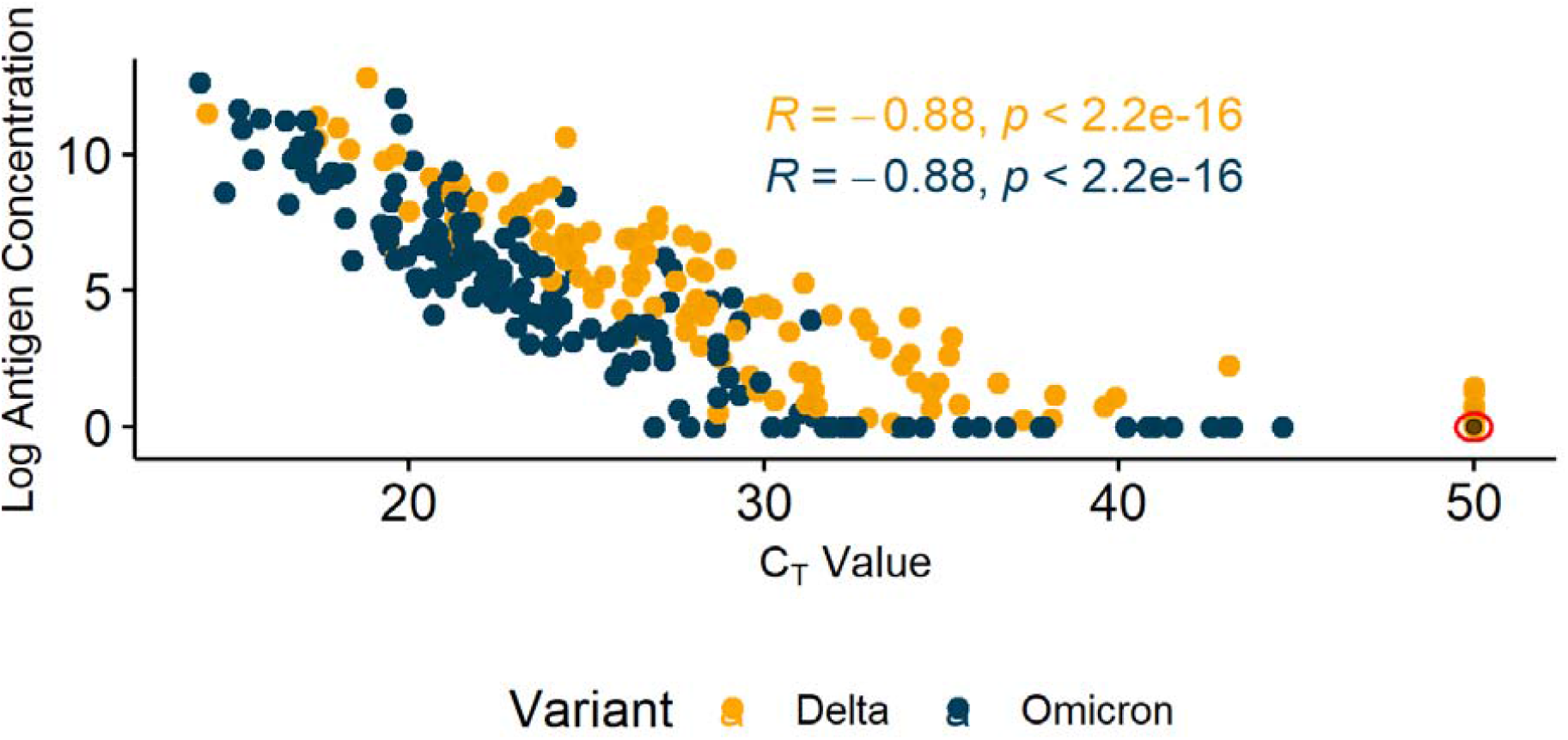
Correlation between antigen concentration and C_T_ value for individual remnant clinical samples. Sequence-verified residual mid turbinate samples from 163 individuals with Delta infection and 169 individuals with BA.1 infection underwent RT-PCR testing using the Xpert Xpress CoV-2/Flu/RSV plus & Xpert Xpress SARS-CoV-2 assays (Cepheid) and protein testing using the Simoa SARS-CoV-2 N Protein Antigen assay (Quanterix), according to the manufacturer’s instructions. Y-axes reflect natural log(n+1) transformed antigen concentration. The red circle signifies 41 Delta samples whose C_T_ values were above the assay detection limit and antigen concentrations were below the assay detection limit. Abbreviations: Cycle threshold (C_T_).

The magnitude of C_T_ difference between Delta and Omicron samples (6.8 cycles, for a given antigen concentration) was greater than the C_T_ difference expected from the different number of freeze-thaws they had undergone (1.9 cycles, with a concomitant decrease in antigen concentration by 16%, Supplementary Material, **Figure S4, Table S2**). Thus, we infer that differential freeze-thaw conditions are likely to explain some, but not all, of the discrepancy we observed. The observation that Omicron samples have a lower amount of antigen-per-RNA also helps to explain our finding that rapid antigen test sensitivity is different when using C_T_ versus antigen concentration as a comparator for RCS pools, all of which underwent the same number of freeze thaws.

### Omicron samples have lower infectivity than Delta samples

Given the observed discrepancy between protein concentration and C_T_ value for Delta and Omicron RCS in this study, we assessed whether there was a difference in virus infectivity from these samples. Seventy-five Delta and 85 Omicron clinical samples with C_T_ <30 were tested against Calu-3 cells in duplicate. Calu-3 cells were infected by 37 (49.3%) of Delta and 37 (43.5%) of Omicron samples (**Figure S5**). ELISpot panels showing representative data are shown in **Figure S6**. Interestingly, infectivity appeared to be inversely associated with C_T_ value for Delta but not Omicron samples (**Figure S5**). We formally assessed this using logistic regression analysis. In univariate analysis, Calu-3 infectivity was inversely associated with C_T_ value for Delta (odds ratio[OR]=0.75, 95% CI: 0.63-0.88) but not Omicron (OR=1.03, 95% CI: 0.90-1.17), and was not associated with antigen concentration, vaccine status, symptom duration, or age (**Table S3**). In multivariate analysis, Calu-3 infectivity remained inversely associated with C_T_ value for Delta (**Table S3**). Specifically, for every 1-cycle increase in C_T_ value, the odds of having a positive Calu-3 result decreased by 28% (95% CI: 14%-42%) for Delta samples. Again, there was no association between Calu-3 infectivity and antigen concentration for either Delta or Omicron samples.

Thus, Omicron samples in this study had less antigen-per-RNA and less infectivity than Delta samples, and there was no association between RNA level (as measured by C_T_) and infectivity for Omicron samples.

## DISCUSSION

Overall, we found that most commercially available RATs had similar sensitivity in detecting Omicron and Delta when antigen concentration was used as a comparator. However, when C_T_ value was used as a comparator, most RATs had a lower sensitivity for Omicron than Delta.

These findings are largely consistent with prior studies showing lower sensitivity of RATs in detecting Omicron than Delta when using C_T_ value as a comparator, especially for samples with low RNA concentration. Osterman *et al*. found a 10-100-fold higher LoD for Omicron compared to Delta among nine RATs in Germany (these tests did not overlap with the tests used in our study) (2). Bayart *et al*. found lower sensitivity for Omicron (0%-23%) than Delta (32%-80%) across 6 RATs in Belgium for clinical samples with C_T_>25 (4). In recent preprints, Bekliz *et al*. found lower sensitivity for Omicron than Delta across 7 RATs (4 of which were statistically significant), and Landaverde *et al*. found low sensitivity of BinaxNOW for detecting Omicron especially with C_T_>23 (5, 6). Only one recent study has shown a high sensitivity of BinaxNOW for Omicron, using clinical samples with C_T_ up to 30 (3). A few other studies reported similar sensitivity for RATs in detecting Omicron and Delta, but these were based on serially diluted, cultured virus, not clinical samples. For example, Deerain *et al*. reported high sensitivity for both variants up to C_T_=25 and essentially no detection at C_T_=28 for 10 RATs (mostly non-overlapping with ours) (7). Stanley *et al*. found decreased sensitivity for Delta compared to Omicron and WA1 (8). All these prior studies used C_T_ value or RNA concentration as a comparator, and none reported antigen concentration. Thus overall, there is accumulating evidence that many RATs demonstrate lower sensitivity for Omicron than Delta for primary clinical samples when using C_T_ value as a comparator, consistent with our findings.

Our study using clinical samples offers a potential explanation for the apparent lower sensitivity of RATs for Omicron, by investigating variant-specific discrepancies in antigen concentration versus C_T_ value. Specifically, the Omicron samples in this study had a lower amount of antigen-per-RNA than the Delta samples, and because RATs detect antigen rather than RNA, they appear less sensitive for Omicron when C_T_ value is used as a comparator. By contrast, when we used antigen concentration as a comparator (an “apples to apples” comparison), we found that most RATs had similar sensitivity for Omicron and Delta.

We considered several technical factors that could account for differences in measurement of RNA and antigen concentration between variants in this study. Whereas Delta clinical samples underwent RNA and protein concentration testing after two freeze-thaw cycles, Omicron clinical samples were tested fresh. Results from our freeze-thaw experiment suggest that this difference in sample handling could account for some, but not all, of the observed variant-specific differences in the ratio between RNA (Ct value) and antigen concentration. Furthermore, RCS pools, which had undergone the same handling conditions for both Delta and Omicron (i.e. samples in both variant pools had the same number of freeze-thaws), also showed a discrepancy in results when antigen versus RNA concentration was used as a comparator. Because our results were consistent across two different RT-PCR assays (Xpert Xpress CoV-2/Flu/RSV *plus* assay (Cepheid) for individual RCS and CDC N2 assay for RCS pools), differences in RT-PCR efficiency are unlikely to account for our findings.

We also considered whether Omicron-specific mutations may have affected the performance of diagnostic antibodies used in these assays. A recent study mapped the N protein epitopes recognized by antibodies in many SARS-CoV-2 antigen tests and identified escape mutations using deep mutational scanning (9). The key amino acid positions identified considering both antibodies used by the Simoa SARS-CoV-2 N Protein Antigen Test (Quanterix) (5, 36-41, 51-53, 56, 62, 66, 71-73, 82-87, 95, 98-101, 108-117, 128-133, 143, 158-161, 167, 171-173) are not canonically mutated in Omicron, and were not specifically mutated in the samples from this study. The same study assessed antibodies for several RATs included here and found no overlap between variant specific mutations and antibody escape mutations. Thus, variant-specific mutations are unlikely to account for differences in the measurement of antigen concentration in this study.

Overall, we infer that the Omicron samples in this study truly had a lower amount of antigen-per-RNA than the Delta samples. There are several potential explanations for this based on viral dynamics over the course of infection. Early studies of SARS-CoV-2 showed that viral load generally peaks around day 3 of viral shedding, just before or at the time of symptom onset, and clears after 7-10 days (10-13). Antigen detection peaks later, generally several days after symptom onset (14), and thus antigen detection frequently lags behind RNA detection (15). Individuals in our study infected with Omicron presented for testing sooner after symptom onset than individuals infected with Delta, and thus our Omicron samples may have been collected at a time when antigen levels may still have lagged behind RNA levels.

In addition, there may be variant-specific differences in viral dynamics, because of either intrinsic biological differences in viral replication and pathogenesis, and/or differences in the characteristics of individuals who are infected with each variant. For example, due to the timing of vaccine booster rollouts, individuals infected with Omicron are likely to have been vaccinated more recently (and with more doses) than individuals infected with Delta. Consistent with this, individuals in our study infected with Omicron had been vaccinated more recently than individuals infected with Delta. In a recent study, boosted individuals infected with Omicron were slower to clear viral RNA than unboosted individuals infected with Delta or Omicron, but the effect on antigen dynamics remains unknown (16). Finally, there may be variant-specific differences in the viral lifecycle that lead to differences in RNA and antigen concentration, such as differential sgRNA transcription, gene expression, protein degradation, or protein aggregation. Compatible with our findings, a recent report demonstrated a decrease in the sensitivity of RATs over time since the start of the pandemic, including during the Omicron era (17). The authors posited that increased immunity, including through vaccination, led to early symptom onset and early testing, before epithelial cell shedding had generated high concentrations of nucleocapsid protein.

Together with these prior studies, our results support a model in which individuals infected with Omicron presented for testing earlier in the course of infection, when antigen concentration lagged behind RNA concentration, leading to an apparent decrease in rapid antigen test sensitivity when C_T_ value is used as a comparator. There are likely multiple factors contributing to the earlier presentation for testing of individuals infected with Omicron, one of which may be a more rapid and robust symptom onset due to recent/boosted vaccination.

Interestingly, we also found that the Omicron samples in this study had lower infectivity than the Delta samples and there was no correlation between C_T_ value and infectivity for Omicron samples. These findings may be explained by recent observations that Omicron cell entry has a greater dependence on receptor-mediated endocytosis than TMPRSS2-mediated spike cleavage and fusion (18) and Omicron replicates less well in TMPRSS2 expressing cells (19)(20).

This study has several limitations. First, due to the necessary logistical constraints of comparing contemporary to banked samples, the individual RCS tested in this study had undergone different handling for Omicron versus Delta samples. This was mitigated to some extent by also testing pooled RCS, and by explicitly testing the effects of freeze-thaw cycles. In addition, while we tested eight commercially-available RATs using pooled RCS, we were only able to test individual RCS against two RATs, given constraints in sample volume. Finally, the marked differences we observed in infectivity between Omicron and Delta samples must be interpreted in light of recent studies showing important variant-specific differences in cell entry and cell biology.

Nevertheless, our results have important implications for clinical practice and public health. First, we show that the choice of comparator assay plays an important role in interpreting the results of sensitivity evaluations for RATs. Future studies will benefit from the use of well-characterized and standardized reference materials to use in assay testing, as well as careful consideration of duration of symptoms at the time of sample collection. Interestingly, based on our findings, the BinaxNOW assay seems to be the most adversely affected by the Omicron variant relative to its performance against Delta variant, which has practical public health implications given its wide use and large market share in the US. By contrast, most commercially-available RATs have similar sensitivity for detecting Omicron and Delta, when antigen concentration is used as a comparator. This reinforces the effectiveness of existing tests, while also emphasizing the point that a negative RDT early in SARS-CoV-2 infection may have low negative predictive value, and RDT testing should be repeated over time. However, within-patient viral dynamics are evolving throughout the pandemic, likely due to changes in both the virus and the host (e.g., vaccination). Further work is needed to investigate the causes and mechanisms of variant-specific differences in RNA concentration, antigen concentration, infectivity, and viral dynamics, particularly as new variants continue to emerge.

## MATERIALS AND METHODS

All methods are described in detail in Supplementary Materials, below are brief descriptions.

### Study Design

We used sequence confirmed Delta and Omicron BA.1 individual and pooled remnant clinical samples to compare the performance of EUA RATs in detecting these two variants. The N protein content, PCR C_T_ values (as a proxy for RNA concentrations) and ability of individual samples to infect cells in *in vitro* infectivity assays were also measured to comprehensively evaluate differences between Delta and BA.1 variants.

### Preparation of Delta and Omicron RCS Pools

As part of the NIH Variant Task Force, in collaboration with participating labs, we obtained low C_T_, sequence-verified Delta and Omicron remnant clinical samples that remained after diagnostic testing. The N2 C_T_ and N protein concentrations in these remnant clinical samples (RCS) were determined at ACME POCT (The Atlanta Center for Microsystems-Engineered Point-of-Care Technologies) as part of our internal quality control (QC). The CDC N2 PCR assay C_T_ value was used as a proxy for RNA concentration (*details in Supplementary Methods*); N protein concentrations were measured by Simoa (*Supplementary Methods*). Between 4-21 low N2 C_T_ RCS with N protein > 4000pg/mL were pooled to generate each Delta and Omicron pool. These pools were serially diluted, N2 C_T_ and N protein quantified, and used to compare eight EUA RATs.

### Collection and Storage of Individual RCS

We utilized a hospital and community-based approach for enrolling eligible COVID-19 symptomatic patients. For samples collected from July to November 2021 (Delta predominant), mid-turbinate (MT) swabs were collected in 1mL saline and frozen at -80 °C. For use in the current study, samples were thawed, 2mLs sterile saline added, frozen, re-thawed, and then analyzed by Cepheid and Quanterix assays. Subsequently, these samples were thawed and utilized for Binax and QuickVue testing and *in vitro* infectivity assays. MT swab samples collected after January 7, 2022 (Omicron predominant) were collected in 3 mL saline, analyzed fresh by Cepheid and Quanterix assays, and frozen at -80 °C. After one freeze thaw, they were used for Binax testing, QuickVue testing and infectivity assays.

### Antigen testing using Quanterix Simoa Assay

Each pool dilution and every clinical sample used in the current study was analyzed for N protein concentration using the Quanterix HD-X Simoa SARS-CoV-2 N Protein Antigen (RUO) assay (Catalog # 103806), according to manufacturer’s instructions.

### PCR testing of remnant clinical samples using Cepheid

All individual remnant clinical samples used for this study underwent PCR testing using the Cepheid GeneXpert Dx Instrument system with either Xpert Xpress CoV-2/Flu/RSV *plus* cartridges (EUA 302-6991, Rev. B., October 2021) or Xpert Xpress SARS-CoV-2 cartridges (EUA 302-3562, Rev. F January 2021) according to manufacturer’s instructions. For the CoV-2/Flu/RSV plus assay, the resulted SARS-CoV-2 Ct value reflects the first of three gene targets (E, N2, or RDRP) to amplify; for the SARS-CoV-2 assay, both E and N2 Ct values are resulted. To determine whether the CT values from the two Xpert assays could be combined for analysis, the laboratory performed a bridging study to confirm that the SARS-CoV-2 assay E target Ct value correlated tightly with the CoV-2 Ct value from the CoV-2/Flu/RSV plus assay. The samples were thawed, split, and run on both assays in parallel according to manufacturer’s instructions **(Supplementary Table S4)**.

### SARS-CoV-2 genome sequencing

All RCS pools and individual RCSs underwent sequencing at ACME POCT, where libraries were generated using SuperScript First Strand Synthesis kit (Thermo Fisher) followed by Swift Amplicon SARS-CoV-2 Research Panel (Swift Biosciences). Illumina MiSeq was used for sequencing, and viralrecon was used for genome assembly.

### Rapid antigen test testing using pools and individual clinical samples

All rapid antigen testing (pool and individual samples) was performed blinded using the direct swab method where sample was spiked onto the swab and manufacturers’ instructions followed for testing. 20µl sample (as described in the IFU for BinaxNOW TM COVID-19 Ag CARD) was used for BinaxNOW, while 50µl was used for all other RATs tested. After completion, results were unblinded.

### Evaluation of infectious SARS-CoV-2 in individual RCS using Calu-3, Vero-TMPRSS-2, and Vero cells

For *in vitro* infectivity studies, 50µl of each individual RCS was used to inoculate (by spinoculation) cells that were 80-90% confluent growing on a 96-well plate. After two hours, sample was removed, and 50µl Opti-MEM and 150µl methycellulose overlay media were added. This portion of the assay was conducted in the BSL3 facility as live lab-propagated SARS-CoV-2 (Delta and BA.1) of known TCID_50/ml_ were used as positive controls. After 3-6 days of incubation (depending on the cell line used), cells were washed with 1XPBS, fixed with chilled 1:1 methanol acetone, permeabilized with 0.2% TritonX, blocked with 1% milk, and then assayed for focus forming units (FFU) by staining with anti-nucleocapsid antibody. Stained foci were read using an ELISpot CTL reader.

### Statistics

Some C_T_ values were above the limit of detection and were therefore set to 50, above the highest recorded C_T_ value. Likewise, some antigen concentrations were below the limit of detection by Simoa and were therefore set equal to zero (0), below the lowest detected antigen concentration. For all analyses, any observations that required imputation were removed in subsequent sensitivity analyses. To meet normality and homoskedasticity assumptions for the linear regression analysis and because there were some values set equal to zero, we used a log(n+1) transformation on antigen concentrations.

We calculated the clinical sensitivity of BinaxNow and QuickVue as well as their corresponding 95% confidence intervals for Delta and Omicron samples overall and by C_T_ or antigen concentration thresholds. Clinical sensitivity was calculated by dividing the number of positive tests by the number of positive participants (samples). The sensitivity of Delta and Omicron samples on the same platform were statistically compared through chi-square or Fisher’s exact tests.

Additionally, we examined and quantified the relationship between C_T_ values and antigen concentration in COVID-19 positive samples from both the Delta and Omicron dominant eras. We calculated Pearson’s correlation coefficient between C_T_ value and antigen concentration, and performed a linear regression analysis, predicting C_T_ value from antigen concentration. In the base model, we controlled for variant status. In the full model, we additionally adjusted for vaccine status and symptom duration. Any individual who was unsure of their vaccine status in any capacity was removed from the appropriate regression analyses.

We also evaluated the association between having a positive result for Calu-3 or Vero-TMPRSS-2 culture and C_T_ value, antigen concentration (pg/mL), symptom duration, vaccine status, and age (years) through unadjusted and adjusted logistic regression analyses. There were no asymptomatic individuals included in this analysis, so symptom duration was treated as a continuous variable to better understand the relationship between days of symptoms and infectivity.

All hypotheses’ tests were 2-sided and a p-value below 0.05 was considered significant. Graphs, correlation calculations, and regression modeling were conducted in R v(4.2.0). The sensitivity of RATs and comparisons between sensitivities were calculated and conducted in R v(4.1.3). Tables were created using the gt and gtsummary package and plots were created with the ggpubr, stringr, and ggplot2 package in R (21-25).

### Study approval

The study protocol was approved by the Emory Institutional Review Board and Children’s Healthcare of Atlanta (IRB#00001082). Written informed consent was received prior to participation.

## Data Availability

All data produced in the present work are contained in the manuscript and supplementary data file.

## Author contributions

*AR and AW are co-first authors, and AR is listed first due to earlier involvement in conceptualization of the study and experiments*.

*WAL and AP are co-senior authors, and AP is listed last due to earlier involvement in data synthesis and manuscript writing*.

*Conceptualization: AR, AW, LB, KM, MG, JAS, WAL, AP*

*Methodology: AR, AW, LB, RP, EF, MG, KM, JAS, WOS, TB, HBB, FF, EW, ML, JFK, PR, ALL, RJ, NRP, EAO, JDR, WAL, AP*

*Investigation: AR, AW, LB, RP, EF, MG, KM, WOS, FF, EO, NRP, WAL, AP*

*Funding: WAL*

*Project administration: AR, MG, AP Supervision: AR, AW, MG, WAL, AP*

*Writing – original draft: AP, AW, AR, LB, EF*

*Writing – review & editing: AP, AW, AR, MG, JKF, FF, NRP*

## Acknowledgments

This work was supported by the National Institute of Biomedical Imaging and Bioengineering under the Atlanta Center for Microsystems Engineered Point-of-Care Technologies (ACME POCT). This work was supported by the NIBIB at the NIH under awards 3U54 EB027690-03S1, 3U54 EB027690-03S2, 3U54 EB027690-04S1 and the National Center for Advancing Translational Sciences of the NIH under award UL1TR002378.

We thank Nils Schoof for providing Delta and BA.1 variant of SARS-CoV-2 and TMPRSS2 cells used in infectivity assays.

We thank Children’s Healthcare of Atlanta and Emory University’s Children’s Clinical and Translational Discovery Core for sample acquisition, selection, processing and storage.

