## Supplementary material for "Sensitivity of Rapid Antigen Tests Against SARS-CoV-2 Omicron and Delta Variants"

#### Supplementary Methods

##### Sample Collection and Storage

The Atlanta Center for Microsystems Engineered Point-of-Care Technologies (ACME-POCT) network utilized hospital and community-based COVID-19 testing centers for enrolling patients. Point of care testing sites included Emory University affiliated hospitals, Grady Memorial Hospital, Children's Hospital of Atlanta, and various community-based drive-through testing centers. Eligible participants were identified consecutively at each study site via review of COVID-19 symptoms. Exclusion criteria included asymptomatic patients, those with symptoms associated with COVID-19 for ≥ 7 days, and unable to provide informed consent. Clinical and demographic variables were collected in a centralized, web-based database (REDCap, Nashville, TN). The study protocol was approved by the Emory Institutional Review Board and Children's Healthcare of Atlanta (IRB#00001082).

For samples collected during July to November 2021, mid-turbinate nasal (MT) swabs were collected using CDC guidelines in 1ml of saline in 1.8ml cryovials and were stored in a 4°C refrigerator until courier pick up (8 hours maximum). Samples were couriered to the Clinical & Translational Discovery Core (CTDC) at the end of the day for long-term storage. All samples are location managed in Nautilus LIMS (Location Information Management System) and stored in temperature-monitored -80°C freezers after arrival at CTDC. For this study, sequence confirmed Delta variant mid-turbinate swabs were thawed and diluted with an additional 2ml saline into a 3.6ml cryovial; the MT swabs were transferred to the 3.6mL cryovial, re-frozen, and couriered to ELIAD on dry ice for further analysis (< 4 hours at 4°C during dilution and transfer). Samples were thawed and underwent analysis by Cepheid and Quanterix Simoa Assay as described below. Samples were again frozen and stored at -80°C before thawing for testing by two lateral flow assays (Abbot BinaxNOW and Quidel QuickVue) as described below.

For samples collected after January 7, 2022, MT swabs were collected using CDC guidelines in 3ml saline in a 3.6 ml cryovial and were stored in a 4°C refrigerator until courier pick up. Swab remained in the sample. Samples were couriered to ELIAD at the end of the day. After arrival at ELIAD, samples were analyzed on Cepheid and by Quanterix Simoa Assay. After analysis and within 24 hours at 4°C, samples were frozen and stored in temperature monitored, -80°C freezers and are location managed in Nautilus LIMS. Samples were thawed and used for LFA (Abbot BinaxNOW and Quidel QuickVue), as described in the next section.

**Antigen testing**

Quantitative SARS-CoV-2 antigen testing of clinical samples and pools was performed on the Quanterix HD-X using the SARS-CoV-2 N Protein Antigen (RUO) assay (Catalog # 103806), according to manufacturer instructions. Briefly, for this 2-step ELISA, 25ml bead reagent conjugated with anti-nucleocapsid antibody was incubated for 35 minutes with 100ml of 4x diluted sample and 20ml detector (biotinylated anti-nucleocapsid antibody). Beads were separated magnetically and washed, then 100ml streptavidin beta-galactosidase (SBG) was added and incubated for 5 minutes. The beads were again magnetically separated and washed, then resuspended in a resorufin ß-D-galactopyranoside (RGP) substrate solution, transferred to a flow cell disc that contained a microarray of femtoliter reaction chambers, and sealed with oil.

Antigen/antibody complexes were detected by fluorescence and counted digitally. For each analysis, an 8-point calibration curve was generated from 2 replicates of 8 calibrators, ranging from 0.228 pg/mL to 192 pg/mL. A four-parameter logistic fit (4PL) was used to generate the curve with $1/y^2$ weighting. For quality control of each run, a negative control, low-, and high-positive controls were measured alongside the samples. Acceptance criteria are as follows: Negative control must be below 3.00 pg/mL. Low- and high- positive measurements should be within 20% of manufacturer specifications. Each run was reviewed for quality by a certified MLS before results were released. Specimens were assayed in duplicate, and in the event a sample was outside the calibrated range of the assay, (~800pg/mL), the sample was diluted 10- or 100-fold and re-tested; final concentrations were calculated by multiplying by the dilution factor.

**PCR testing of clinical samples**

PCR testing of individual clinical samples was performed using the Cepheid GeneXpert Dx Instrument System with Xpert Xpress CoV-2/Flu/RSV *plus* cartridges (EUA 302-6991, Rev. B., October 2021). Testing was performed as described by manufacturer with new lots or shipments assayed in parallel. This test detects and differentiates RNA from SARS-CoV-2, Influenza A, Influenza B, and Respiratory Syncytial Virus (RSV) using multiple targets. Only results from SARS-CoV-2 are included herein. The reported PCR cycle threshold ($C_T$) value represents the first target to amplify from one of the three unique SARS-CoV-2 genome target sequences: nucleocapsid (N), envelope (E), and RNA-dependent RNA polymerase (RdRP). A Sample Processing Control (SPC) is also included to monitor for inhibitors of the RT-PCR reaction and to assess appropriate sample processing. Interpretations are generated automatically using algorithms within the GeneXpert System Software. Patient samples tested before 01/07/2022 used Xpert Xpress SARS-CoV-2 cartridges (EUA 302-3562, Rev. F January 2021*)* in accordance with manufacturer's instructions. This test detects SARS-CoV-2 RNA and generates Ct values for both the E and N2 targets. The laboratory performed a bridging study to confirm that the E target was the appropriate comparator with the Xpert Xpress CoV-2/Flu/RSV plus results (**Table S4**).

**Preparation of Delta and BA.1 pools and dilutions for testing LFA sensitivity**

Sequence verified non-heat-inactivated (live) samples that remained after clinical testing (remnant clinical samples, RCS) were selected from diagnostic labs partnering with the NIH Variant Task Force and shipped to Emory University on dry ice. Samples for pooling were selected based on low N2 Ct (<24 as determined by the 2019-nCoV CDC EUA assay), N gene sequence compatible with Delta and BA.1 variants and having >4000pg/ml N protein as determined by the Quanterix assay described in the previous section. The number of RCS used for each pool varied between 4 and 21 depending on the volume of each individual sample and the final pool volume needed for testing. For preparing Delta pool 1, we pooled 4 RCS. After this pool was completely utilized, we used the same original stock to prepare pool 1.1 for additional testing. For testing with BA.1 we used a total of 3 pools, making a new pool each time the previous pool was used up. Pool 1 utilized 6 BA.1 samples, while Pool 2 used 18 and Pool 3 used 21 samples. After the pool was prepared, RNA was extracted from 140µl undiluted pool using MagMax Viral RNA Isolation Kit (Applied Biosystems) in a KingFisher Apex system (Thermo Fisher Scientific). RNA was eluted in 60µl and 5 µl of the eluate was used in a one-step RT-PCR reaction and reverse transcribed into cDNA with qScript XLT 1-Step RTqPCR ToughMix (QuantaBio) using 2019-nCoV CDC EUA Kit, 1000 reaction combined Primer/Probe Mix (N2 gene) (IDT, Catalog No. 10006770) in a LightCycler 480 II instrument (Roche) or ddPCR (BioRAD). The N2 Ct concentration of each undiluted pool was determined using the CDC N2 assay. After determining the N2 Ct concentration of the pool, it was serially diluted in SARS-CoV-2 negative Lee Biosolutions Nasal Wash (Catalog number 991-26-P-1) such that N2 Ct of dilutions ranged from ~18-31. Subsequently, each RNA stock was sequenced as described below to confirm the variant. The N protein concentration of each dilution of every pool was determined using the Quanterix Simoa Assay, as described later.

**SARS-CoV-2 genome sequencing**

We analyzed SARS-CoV-2 genome sequences from 72 individual Delta and 152 individual Omicron samples, as well as the RCS pools. For both clinical samples and pools, SARS-CoV-2 sequencing libraries were generated using the SuperScript IV First Strand Synthesis kit (Thermo Fisher) followed by the Swift Amplicon SARS-CoV-2 Research Panel (Swift Biosciences), as previously described (*26*). Sequencing was performed with paired-end 150bp reads on an Illumina MiSeq. The consensus SARS-CoV-2 genome was assembled using viralrecon version 2.4.1 with reference sequence MN908947.3 (*27*). Variants were classified by Pangolin (*28*). All sequence data is available via GISAID (Accession numbers listed in Supplementary Data File, Column X) and NCBI under BioProject PRJNA634356. Specific N protein mutations identified in samples used in this study are depicted in Supplementary Data File, Column Y.

**LFA testing**

Clinical samples and serially diluted pools were used to test each LFA according to the IFU supplied with the test kit. For all, the direct swab method was used, in which 20mL (for BinaxNOW) or 50mL (for all other LFAs) of sample was spiked onto the swab, and then the IFU followed. All testing was done in a blinded manner, and after completion of testing, tests results were unblinded and data interpreted.

**Infectivity assays**

Samples: SARS-CoV-2 mid-turbinate swab samples (in saline) from individuals with COVID-19 were received from Emory Clinic and confirmed by sequencing: Delta (77 samples) and Omicron (87 samples) variants. Samples were handled in the biosafety level (BSL2*) and the (BLS3) laboratories at Emory University.

Viral agents (kindly provided by Nils Schoof): The SARS-CoV-2 Delta variant propagated at ACME/Emory (virus stock#13 at $2.2x10^6$ FFU/mL or $3.1x10^6$ $TCID_{50}$/ml) was used as a positive control for the plates containing Delta clinical samples, and the Omicron (BA.1) variant propagated at ACME/Emory (virus stock#53 at $2.5x10^6$ FFU/ml or $3.5x10^6$ $TCID_{50}$/ml) was used as a positive control for the plates containing Omicron clinical samples.

Cell types: Three cell types were used for infectivity assays, including Vero E6 cells (ATCC), and VeroE6/ Type 2 transmembrane serine protease TMPRSS2 (kindly provided by Nils Schoof), and Calu-3 (ATCC). TMPRSS2-overexpressing Vero E6 cells were used for enhancing SARS-CoV-2 entry into host cells (*19, 29*)

All three cell types were maintained in the appropriate medium supplemented with 5-10% fetal bovine serum (FBS) and antibiotics (100 units/ml penicillium/100 µg/ml streptomycin (Pen/Strep), and 1% L-glutamine), including:

1. Vero E6 cells: Minimum Essential Medium (MEM) supplemented with 5% FBS (MEM-5).
2. VeroE6/TMPRSS2 cells: Eagle's Minimum Essential Medium (EMEM) supplemented with 10% FBS and 10µg/ml puromycin.
3. Calu-3 cells: Dulbecco's Modified Eagle Medium/Nutrient mixture F-12 (DMEM/F-12) supplemented with 10% FBS.
4. Overlay medium: OptiMEM + 2%FBS + 2.5 µg/mL amphotericin B (Sigma #A2942-20mL) + 20 µg/mL Cipro (Corning #61-277-RF 1g) + 2% methylcellulose (Sigma #M0512-2506)
5. Primary antibody (SARS-CoV/SARS-CoV-2 Nucleocapsid Antibody, Rabbit Mab) (Sino Biological Cat# 40143-R-001)
6. Secondary antibody (HRP-Goat Anti-Rabbit) (Invitrogen, Cat#656120)
7. Pierce Clear Milk Blocking Buffer (10X) - ThermoFisher
8. TrueBlue Peroxidase Substrate - SeraCare (KPL #50-78-02)

**Focus-Forming Assay (FFA) (*30, 31*):** Vero-E6 (5,000 cells/well), VeroE6-TMPRSS2 (10,000 cells/well), or Calu-3 (50,000 cells/well) cells were seeded in 96-well plates and incubated at 37$^{o}$C, 5% $CO_2$. The next day or until the cells reached ~80-90% confluency, the medium was removed from each plate. The cells were inoculated with 50µL of undiluted clinical samples for two h (one h spinoculation at 918xg followed by one h incubation at 37$^{0}$C, 5% $CO_2$). The inoculum was removed from cells, followed by adding 50µl Opti-MEM medium (ThermoFisher) and 150µL methylcellulose overlay medium; This work was performed in the BSL3 facility. The SARS-CoV-2 Delta variant (stock#13) was used as a positive control for the plates containing Delta clinical samples, and the Omicron (virus stock#53) was used as a positive control for the plates containing

Omicron clinical samples. The two viral stocks were diluted at 1:100, 1:1,000, 1:10,000, or 1:100,000 in OptiMEM medium and inoculated to the cells for 1-2 h at 37°C, 5% $CO_2$. The negative controls (cells in OptiMEM) were included in each plate. All samples were performed in singles and positive and negative controls in duplicates. A total of four independent experiments were performed for Delta and Omicron samples: Two for Calu-3, one for Vero E6, and one for VeroE6/TMPRSS2). After infection, the inoculum was removed from the cells and replenished with 50µL OptiMEM and 150 µL overlay media for 3-6 days. After six days of incubation (~3-4 days incubation with the positive controls), cells were washed three times with 1x phosphate-buffered solution (PBS), fixed with a 1:1 ratio of chilled methanol and acetone at room temperature for 30 minutes, washed once with 1xPBS, permeabilized with 0.2% Triton X-100 at room temperature for 10 minutes. Cells were washed once with 1xPBS and blocked with 1x milk for at least 20 minutes at room temperature. Plates were then transferred to the BSL2*, and cells were incubated with primary antibody nucleocapsid antibody at 1:5,000 in 1% milk at 37°C for two h (TMPRSS2) or ~18 h (Vero E6 or Calu-3). After two h (for TMPRSS2) or the next day (for Vero E6 or Calu-3), cells were washed twice with 1xPBS and incubated at 37°C for one h with the secondary antibody at 1:5,000 in 1% milk. Cells were washed twice with 1xPBS, dried at room temperature for 5-10 minutes, followed by the addition of TrueBlue HRP substrate, and incubated at room temperature on a rocker for up to one hour. After removing the substrate, cells were dried, and the stained foci were visualized and imaged using an ELISpot reader (CTL). Virus infectivity was recorded as the presence of focus-forming units (FFU).

**Foci-forming units (FFA) results and ELISpot reading**

Four independent experiments were performed: All Delta and Omicron clinical samples were performed twice in Calu-3 and once in Vero E6 and VeroE6/TMPRSS2 cells. Clinical samples were performed in a single in each of the four experiments, and negative or positive controls were performed in replicates of 2.

#### Supplementary Results

##### Rapid antigen test sensitivities for Delta and Omicron using serially diluted, pooled clinical samples are similar when using antigen concentration as the comparator, but not when using RNA measured by cycle threshold ($C_T$) value as the comparator.

In addition to LoD comparison, rapid antigen test results were compared across the full spectrum of RNA and antigen concentrations using binary logistic regression. This analysis also showed comparable detection of Omicron and Delta when antigen concentration was used as a comparator (**Figure S2**) but poorer detection of Omicron than Delta for samples with $C_T$ values in the 23-30 range (**Figure S2**). Because these experiments were performed with serial dilutions rather than independent samples, formal statistical analysis was not undertaken.

##### Omicron samples have lower antigen-per-RNA than Delta samples, even after accounting for freeze-thaw differences

Because the banked Delta RCS had undergone two freeze-thaw cycles prior to the start of this study, whereas the fresh Omicron RCS had undergone none, we evaluated the effect of freeze-thaw cycles on RNA and protein stability, using a different set of 16 samples tested serially before and after freeze-thaw (**Figure S6**, **Table S1**). After two freeze-thaw cycles, $C_T$ increased by a median of 1.9 cycles (maximum 4.6) and antigen concentration decreased by a median of 16%, but with 90-100% loss in two samples. Thus, we inferred that the Delta samples in this study might be expected to have a 4-10-fold reduction in RNA and an approximately 20% reduction in antigen concentration, due to the effect of freeze-thaw cycles.

##### Omicron samples have lower infectivity than Delta samples

Infectivity of the same 75 Delta and 85 Omicron samples was also tested in Vero-TMPRSS2 cells (in singlet), and Vero cells (in singlet). Vero-TMPRSS2 cells were infected by 61 (81.3%) of Delta and 33 (38.8%) of Omicron samples, and Vero cells by 17 (22.6%) of Delta and 10 (11.7%) of Omicron samples (**Figure S5**). Similar to our findings with Calu-3 cells, there was a significant association between Vero-TMPRSS2 infectivity and $C_T$ for Delta samples (adjusted odds ratio [aOR]=0.70, 95% CI: 0.49-0.92) but not Omicron samples (aOR=1.11, 95% CI: 0.94-1.33), in both univariate and multivariate (**Table S4**) analysis.

### Supplementary Figures

| Variant / Pool # / Dilution # | Average N2-Ct | Standard Deviation N2-Ct | Average Quanterix (pg/ml) N protein | Standard Deviation Quanterix N protein (n=2) | Binax [n=2] (Lot 1) | ACON [n=2] (Lot 1) | iHealth [n=2] (Lot 1) | Osang [n=5] (Lot 1) | Roche [n=2] (Lot 1) | Roche [n=2] (Lot 2) | Siemens [n=2] (Lot 1) | Siemens [n=2] (Lot 2) | Quickvue [n=5] (Lot 1) |
|---|---|---|---|---|---|---|---|---|---|---|---|---|---|
| Delta Variant / Pool 1 / Dilution 1 | 19.0 | 0.08 | 23425 | 1405 | 100 | 100 | 100 | 100 | 100 | 100 | 100 | 100 | 100 |
| Delta Variant / Pool 1 / Dilution 2 | 20.1 | 0.09 | 11996 | 401 | 100 | 100 | 100 | 100 | 100 | 100 | 100 | 100 | 100 |
| Delta Variant / Pool 1 / Dilution 3 | 21.5 | 0.05 | 3591 | 75 | 100 | 100 | 100 | 100 | 100 | 100 | 100 | 100 | 100 |
| Delta Variant / Pool 1 / Dilution 4 | 22.7 | 0.03 | 2759 | 151 | 100 | 100 | 100 | 100 | 100 | 100 | 100 | 100 | 100 |
| Delta Variant / Pool 1 / Dilution 5 | 24.1 | 0.04 | 887 | 82 | 100 | 100 | 100 | 60 | 100 | 100 | 100 | 100 | 0 |
| Delta Variant / Pool 1 / Dilution 6 | 25.6 | 0.17 | 379 | 3.5 | 100 | 0 | 0 | 0 | 100 | 100 | 100 | 0 | 0 |
| Delta Variant / Pool 1 / Dilution 7 | 26.7 | 0.26 | 148 | 1.8 | 100 | 0 | 0 | 0 | 100 | 100 | 0 | 0 | 0 |
| Delta Variant / Pool 1 / Dilution 8 | 28.1 | 0.06 | 62 | 0.3 | 100 | 0 | 0 | 0 | 0 | 0 | 0 | 0 | 0 |
| Delta Variant / Pool 1 / Dilution 9 | 28.9 | 0.02 | 18 | 2.1 | 0 | 0 | 0 | 0 | 0 | 0 | 0 | 0 | 0 |
| Delta Variant / Pool 1 / Dilution 10 | 30.0 | 0.05 | 12 | 0.8 | 0 | 0 | 0 | 0 | 0 | 0 | 0 | 0 | 0 |
| Delta Variant / Pool 1 / Dilution 11 | 30.9 | 0.08 | 5 | 5 | 0 | 0 | 0 | 0 | 0 | 0 | 0 | 0 | 0 |

| Variant / Pool # / Dilution # | Average N2-Ct | Standard Deviation N2-Ct | Average Quanterix (pg/ml) N protein | Standard Deviation Quanterix N protein (n=2) | Binax [n=5] (Lot 4) | Osang [n=5] (Lot 2) | Quickvue [n=5] (Lot 1) | Boson [n=5] (Lot 1) | Boson [n=5] (Lot 2) |
|---|---|---|---|---|---|---|---|---|---|
| Delta Variant / Pool 1.1 / Dilution 1 | 20.8 | 0.04 | 20861.5 | 491.2 | 100 | 100 | 100 | 100 | 100 |
| Delta Variant / Pool 1.1 / Dilution 2 | 21.5 | 0.09 | 10241.4 | 629.6 | 100 | 100 | 100 | 100 | 100 |
| Delta Variant / Pool 1.1 / Dilution 3 | 22.7 | 0.03 | 4930.1 | 193.8 | 100 | 100 | 100 | 100 | 100 |
| Delta Variant / Pool 1.1 / Dilution 4 | 23.6 | 0.15 | 2887.6 | 82.4 | 100 | 100 | 100 | 100 | 100 |
| Delta Variant / Pool 1.1 / Dilution 5 | 24.2 | 0.04 | 1209.8 | 50.6 | 100 | 100 | 0 | 100 | 100 |
| Delta Variant / Pool 1.1 / Dilution 6 | 26.1 | 0.10 | 756.4 | 27.3 | 100 | 100 | 0 | 100 | 100 |
| Delta Variant / Pool 1.1 / Dilution 7 | 26.8 | 0.15 | 342.9 | 44.7 | 100 | 0 | 0 | 100 | 100 |
| Delta Variant / Pool 1.1 / Dilution 8 | 27.7 | 0.12 | 274.4 | 21.8 | 0 | 0 | 0 | 100 | 100 |
| Delta Variant / Pool 1.1 / Dilution 9 | 28.5 | 0.15 | 114.6 | 3.5 | 0 | 0 | 0 | 0 | 0 |
| Delta Variant / Pool 1.1 / Dilution 10 | 28.9 | 0.03 | 68.4 | 2.0 | 0 | 0 | 0 | 0 | 0 |
| Delta Variant / Pool 1.1 / Dilution 11 | 30.4 | 0.11 | 29.3 | 2.0 | 0 | 0 | 0 | 0 | 0 |

| Variant / Pool # / Dilution # | Average N2-Ct | Standard Deviation N2-Ct | Average Quanterix (pg/ml) N protein | Standard Deviation Quanterix N protein (n=2) | Binax [n=5] (Lot 2) | ACON [n=5] (Lot 2) | iHealth [n=5] (Lot 2) | Roche [n=5] (Lot 1) | Siemens [n=5] (Lot 3) | Quickvue [n=5] (Lot 2) |
|---|---|---|---|---|---|---|---|---|---|---|
| BA.1 Variant / Pool 1 / Dilution 1 | 19.3 | 0.03 | 4100.2 | 110 | 100 | 100 | 100 | 100 | 100 | 100 |
| BA.1 Variant / Pool 1 / Dilution 2 | 19.8 | 0.01 | 1855.4 | 80 | 100 | 100 | 100 | 100 | 100 | 100 |
| BA.1 Variant / Pool 1 / Dilution 3 | 20.9 | 0.05 | 1113.3 | 28 | 100 | 100 | 100 | 100 | 100 | 100 |
| BA.1 Variant / Pool 1 / Dilution 4 | 21.6 | 0.03 | 531.2 | 26 | 100 | 100 | 100 | 100 | 100 | 100 |
| BA.1 Variant / Pool 1 / Dilution 5 | 22.9 | 0.06 | 446.2 | 19.5 | 40 | 100 | 60 | 100 | 0 | 80 |
| BA.1 Variant / Pool 1 / Dilution 6 | 23.9 | 0.01 | 248.5 | 5.4 | 0 | 100 | 0 | 100 | 0 | 0 |
| BA.1 Variant / Pool 1 / Dilution 7 | 24.9 | 0.05 | 134.7 | 3.1 | 0 | 60 | 0 | 80 | 0 | 0 |
| BA.1 Variant / Pool 1 / Dilution 8 | 26.0 | 0.02 | 62.4 | 1.7 | 40 | 0 | 0 | 0 | 0 | 0 |
| BA.1 Variant / Pool 1 / Dilution 9 | 26.9 | 0.07 | 33.5 | 1.4 | 0 | 0 | 0 | 0 | 0 | 0 |
| BA.1 Variant / Pool 1 / Dilution 10 | 27.7 | 0.04 | 17.6 | 0.5 | 0 | 0 | 0 | 0 | 0 | 0 |
| BA.1 Variant / Pool 1 / Dilution 11 | 28.8 | 0.08 | 8.7 | 0.2 | 0 | 0 | 0 | 0 | 0 | 0 |

| Variant / Pool # / Dilution # | Average N2-Ct | Standard Deviation N2-Ct | Average Quanterix (pg/ml) N protein | Standard Deviation Quanterix N protein (n=2) | Binax [n=5] (Lot 2) | Osang [n=5] (Lot 2) | QuickVue [n=5] (Lot 3) | Boson [n=5] (Lot 1) |
|---|---|---|---|---|---|---|---|---|
| BA.1 Variant / Pool 2 / Dilution 1 | 19.8 | 0.10 | 20408 | 82.9 | 100 | 100 | 100 | 100 |
| BA.1 Variant / Pool 2 / Dilution 2 | 20.8 | 0.15 | 11602 | 150.1 | 100 | 100 | 100 | 100 |
| BA.1 Variant / Pool 2 / Dilution 3 | 21.5 | 0.25 | 5209 | 446.1 | 100 | 100 | 100 | 100 |
| BA.1 Variant / Pool 2 / Dilution 4 | 22.7 | 0.06 | 2749 | 267.3 | 100 | 100 | 100 | 100 |
| BA.1 Variant / Pool 2 / Dilution 5 | 23.6 | 0.10 | 1441 | 37.3 | 100 | 100 | 100 | 100 |
| BA.1 Variant / Pool 2 / Dilution 6 | 24.0 | 0.21 | 569 | 30.9 | 100 | 0 | 0 | 100 |
| BA.1 Variant / Pool 2 / Dilution 7 | 24.8 | 0.20 | 326 | 2.5 | 100 | 0 | 0 | 100 |
| BA.1 Variant / Pool 2 / Dilution 8 | 25.8 | 0.06 | 150 | 9.8 | 0 | 0 | 0 | 0 |
| BA.1 Variant / Pool 2 / Dilution 9 | 27.3 | 0.25 | 71 | 2.3 | 0 | 0 | 0 | 0 |
| BA.1 Variant / Pool 2 / Dilution 10 | 28.1 | 0.21 | 37 | 1.7 | 0 | 0 | 0 | 0 |
| BA.1 Variant / Pool 2 / Dilution 11 | 29.1 | 0.15 | 24 | 0.8 | 0 | 0 | 0 | 0 |

| Variant / Pool # / Dilution # | Average N2-Ct | Standard Deviation N2-Ct | Average Quanterix (pg/ml) N protein | Standard Deviation Quanterix N protein (n=2) | Binax [n=5] (Lot 3) | Binax [n=5] (Lot 4) | ACON [n=5] (Lot 2) | QuickVue [n=5] (Lot 3) |
|---|---|---|---|---|---|---|---|---|
| BA.1 Variant / Pool 3 / Dilution 1 | 18.6 | 0.31 | 117088 | 8773.2 | 100 | 100 | 100 | 100 |
| BA.1 Variant / Pool 3 / Dilution 2 | 19.5 | 0.17 | 53210 | 680.1 | 100 | 100 | 100 | 100 |
| BA.1 Variant / Pool 3 / Dilution 3 | 20.7 | 0.04 | 29070 | 550 | 100 | 100 | 100 | 100 |
| BA.1 Variant / Pool 3 / Dilution 4 | 21.5 | 0.14 | 15070 | 641.9 | 100 | 100 | 100 | 100 |
| BA.1 Variant / Pool 3 / Dilution 5 | 22.6 | 0.15 | 6599 | 190.3 | 100 | 100 | 100 | 100 |
| BA.1 Variant / Pool 3 / Dilution 6 | 24.0 | 0.30 | 3499 | 41.4 | 100 | 100 | 100 | 0 |
| BA.1 Variant / Pool 3 / Dilution 7 | 24.7 | 0.10 | 1724 | 72.6 | 100 | 100 | 100 | 0 |
| BA.1 Variant / Pool 3 / Dilution 8 | 25.7 | 0.25 | 891 | 20.6 | 100 | 100 | 100 | 0 |
| BA.1 Variant / Pool 3 / Dilution 9 | 26.7 | 0.09 | 396 | 13 | 80 | 0 | 0 | 0 |
| BA.1 Variant / Pool 3 / Dilution 10 | 27.7 | 0.13 | 183 | 7.1 | 0 | 0 | 0 | 0 |
| BA.1 Variant / Pool 3 / Dilution 11 | 28.4 | 0.36 | 107 | 3.3 | 0 | 0 | 0 | 0 |
| BA.1 Variant / Pool 3 / Dilution 12 | 29.5 | 0.11 | 48 | 2.7 | 0 | 0 | 0 | 0 |

**Fig. S1. Complete results of LFA testing using serially diluted pools of Delta and BA.1 samples.** Values shaded in green indicate 100% (5/5) positive results. Values highlighted in peach indicate numbers that were between 0% and 100% positive.

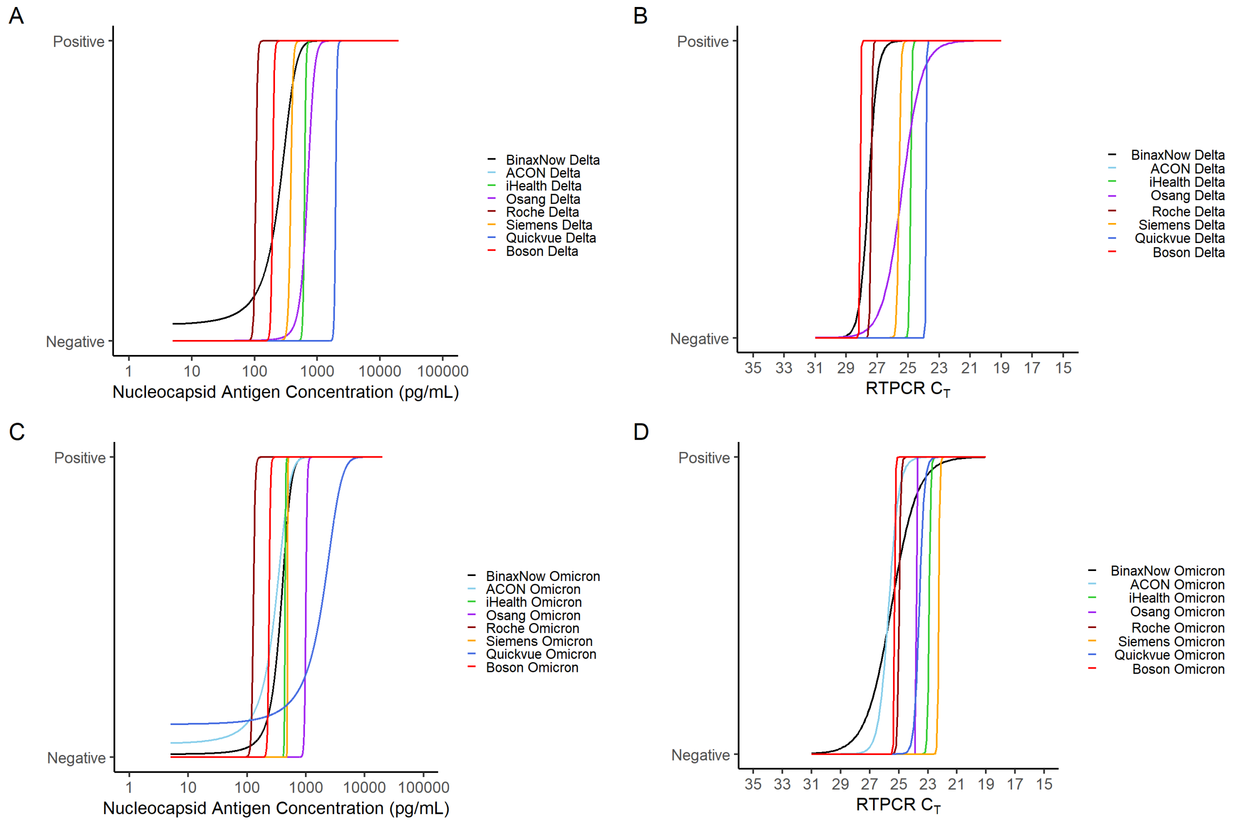

**Fig. S2. Logistic regression results of testing 8 commercially available rapid antigen tests against remnant clinical sample pools.** Plots of binary logistic regressions, in which the outcome is positive rapid antigen test, and the predictor is antigen concentration **(A, C)** or $C_T$ value **(B, D)** for Delta (A-B) and Omicron (C-D) RCS pools. Abbreviations: Cycle threshold ($C_T$), Center for Disease Control (CDC)

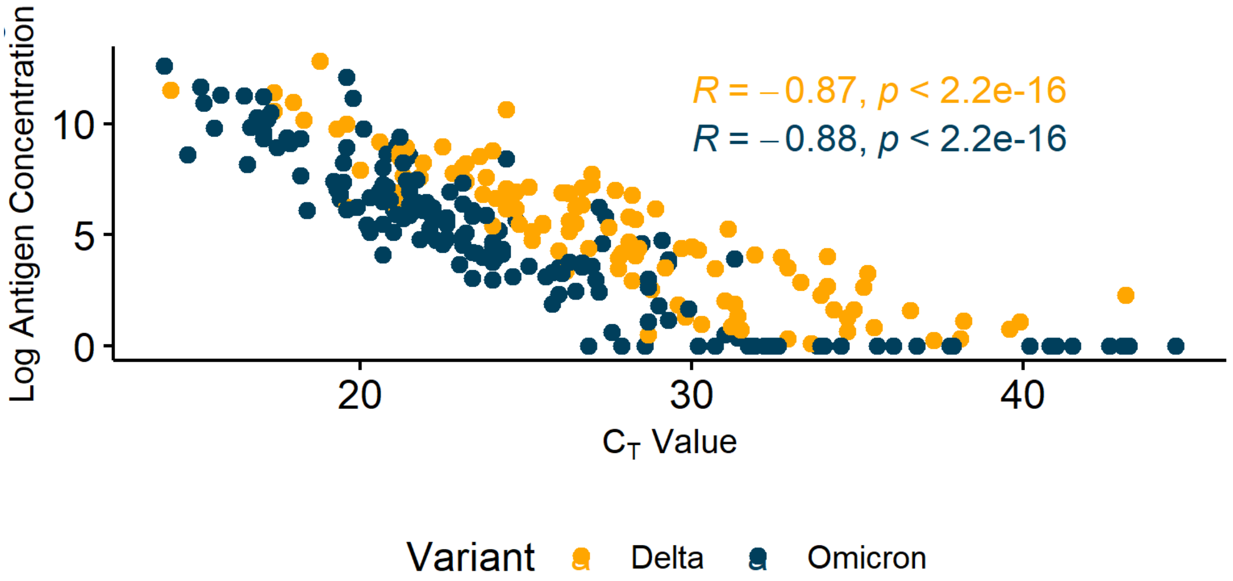

**Fig. S3**. **Scatterplots of $C_T$ value with natural log(n+1) transformed antigen concentration.** Scatterplot demonstrating the overall correlation of $C_T$ value with natural log(n+1) transformed antigen concentration after removing observations originally above or below the detectable limit.

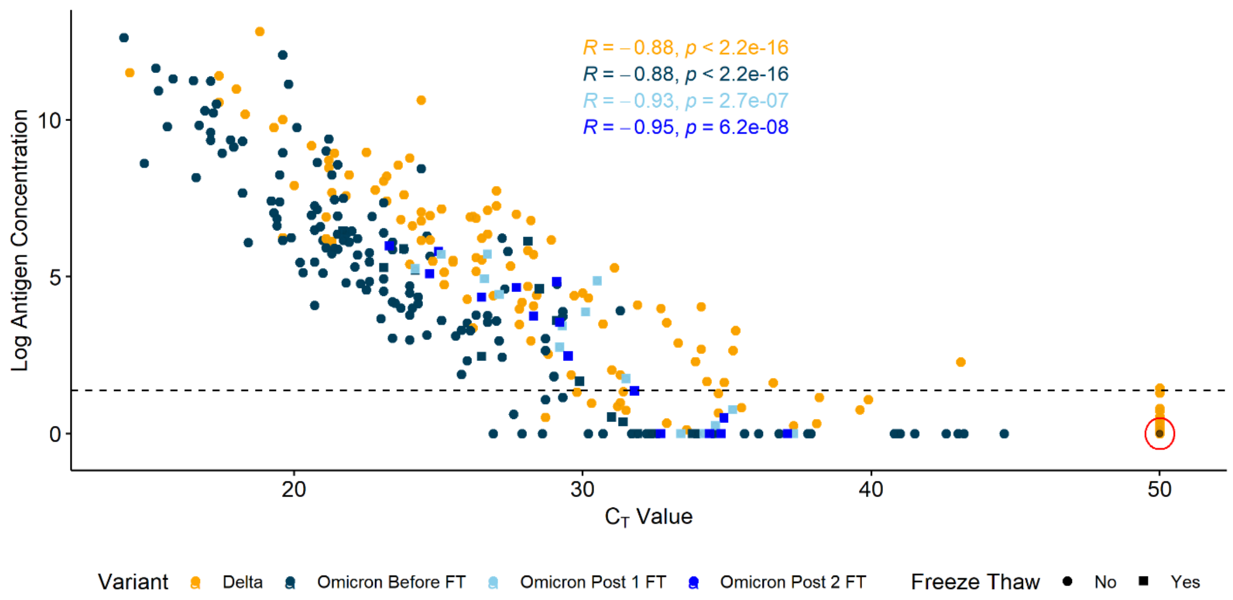

**Fig. S4**. **Sixteen randomly selected Omicron samples with equidistant $C_T$ values were subjected to two freeze thaws.** After each freeze thaw, the $C_T$ value and antigen concentration was recorded. The dashed cutoff line represents the antigen LOD cutoff at 3 pg/mL. As in Figure 3, the red circle signifies 41 Delta samples with $C_T$ values above calibration and antigen concentrations below calibration.

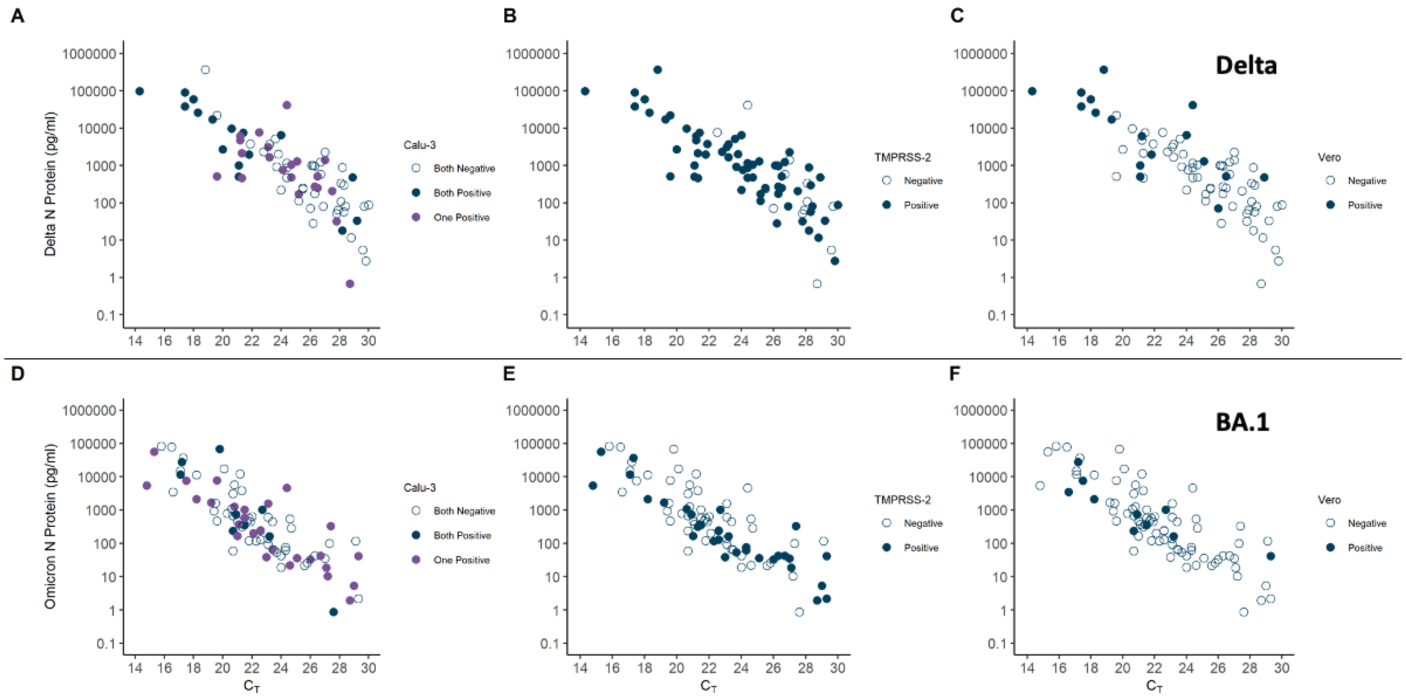

**Fig. S5. Infectivity of Delta (A, B, C) and Omicron (D, E, F) remnant clinical samples in three different cell types: Calu-3 (A, D), Vero-TMPRSS2 (B, E), and Vero (C, F).** Scatterplots represent antigen concentration versus $C_T$ for individual remnant clinical samples; samples with negative infectivity for a given cell type are empty and samples with positive infectivity are filled. Abbreviations: Cycle threshold ($C_T$).

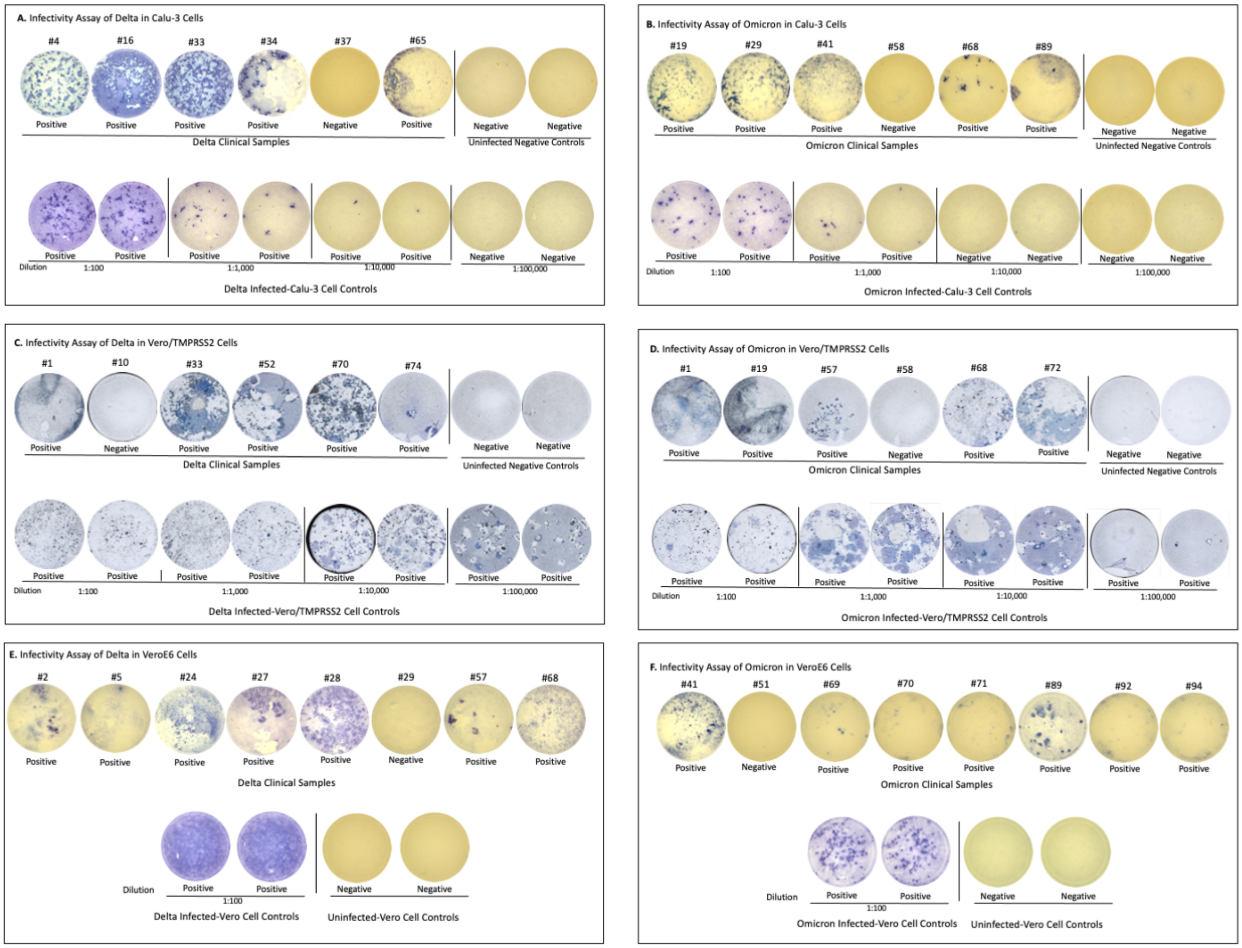

**Fig. S6**. **Representative panels showing Focus-forming infectivity assay of clinical samples from individuals with COVID-19 Delta or Omicron variants of SARS-CoV-2 using an ELISpot reader.** Clinical samples or uninfected negative control were added to the cells and spinoculated for 1 hr at 918xg followed by 1 hr incubation at 37oC, 5% CO2. Two hr after inoculation, the inoculum was removed from the cells, followed by adding 50 µL OptiMEM and 150 µL methylcellulose overlay medium. The SARS-CoV-2 Delta variant (stock#13) was used as a positive control for the plates containing Delta clinical samples, and the Omicron (virus stock#53) was used as a positive control for the plates containing Omicron clinical samples. The two viral stocks were diluted at 1:100, 1:1,000, 1:10,000, or 1:100,000 in OptiMEM medium and inoculated (50 µL) to the cells for 1-2 hr at 37oC, 5% CO2. For VeroE6 cell infection, viral stocks were diluted at 1:100 in OptiMEM. After infection, the inoculum was removed from the cells and replenished with 50µL OptiMEM and 150 µL overlay media for 3-6 days. All clinical samples were performed in singles undiluted. Controls were performed in duplicates. Experiment performed with Delta samples in **(A)** Calu-3 **(C)** VeroE6/TMPRSS2 **(E)** VeroE6 cells, and Experiment performed with Omicron samples in **(B)** Calu-3 cells **(D)** VeroE6/TMPRSS2 and **(F)** VeroE6 cells.

**Supplementary Tables**

**Table S1**. **Association between $C_T$ value and natural log(n+1) transformed antigen concentration, variant, vaccine status, and symptom duration for 104 Delta and 165 Omicron samples, excluding samples with above calibration $C_T$ value or below calibration antigen concentration.** No asymptomatic patients were present in this analysis, so symptom duration is a continuous variable.

| | Base Model | | | Full Model | | |
|---|---|---|---|---|---|---|
| **Variable** | **Beta** | **SE** | **p-value** | **Beta** | **SE** | **p-value** |
| Log (Antigen Concentration + 1) | -1.7 | 0.057 | <0.001 | -1.6 | 0.057 | <0.001 |
| Variant | | | | | | |
| Delta | Ref | — | | Ref | — | |
| Omicron | -3.0 | 0.377 | <0.001 | -2.8 | 0.451 | <0.001 |
| Vaccine Status | | | | | | |
| Not Fully Vaccinated | | | | Ref | — | |
| At Least Fully Vaccinated | | | | 0.30 | 0.862 | 0.7 |
| Symptom Duration | | | | 0.13 | 0.060 | 0.04 |
| Days Since Last Vaccine | | | | | | |
| Unvaccinated | | | | Ref | — | |
| Within the Last 90 Days | | | | 0.25 | 0.833 | 0.8 |
| Between 91 and 180 Days Ago | | | | -0.70 | 1.01 | 0.5 |
| Between 181 and 270 Days Ago | | | | -1.1 | 0.98 | 0.3 |
| More than 270 Days Ago | | | | -0.13 | 1.23 | >0.9 |

SE = Standard Error, Ref = Reference Level

NOTE: Beta coefficients have been rounded but percentage change calculations were computed before rounding and therefore may be different.

**Table S2. Sixteen randomly selected Omicron samples with equidistant CT values were subjected to two freeze thaws.** After each freeze thaw, the CT value and antigen concentration was recorded. All samples were thawed and re-tested on 7/13/2022 (FT1) and 7/14/2022 (FT2).

| ID | Lineage | Date Collected | Original Cepheid $C_T$ | Original HD-X Ag Conc. (pg/mL) | Retest Cepheid $C_T$ (FT1) | Retest HD-X Ag Conc. (pg/mL) (FT1) | Retest Cepheid $C_T$ (FT2) | Retest HD-X Ag Conc. (pg/mL) (FT2) |
|---|---|---|---|---|---|---|---|---|
| F1 | BA.1 | Jan 2022 | 21.7 | 638 | 25.1 | 301 | 23.3 | 396 |
| F2 | BA.1 | Jan 2022 | 21.8 | 625 | 24.2 | 191 | 24.7 | 163 |
| F3 | BA.1 | Dec 2021 | 23.1 | 198 | 26.6 | 138 | 27.7 | 104 |
| F4 | BA.1 | Jan 2022 | 23.8 | 358 | 27.1 | 83.7 | 26.5 | 76.5 |
| F5 | BA.1.1 | Jan 2022 | 24.2 | 182 | 26.7 | 305 | 25 | 331 |
| F6 | BA.1.1 | Jan 2022 | 26.5 | 10.8 | 29.3 | 30.1 | 29.2 | 33.8 |
| F7 | BA.1.1 | Feb 2022 | 28.1 | 461 | 30.1 | 47.3 | 28.3 | 41.1 |
| F8 | BA.1.1 | Jan 2022 | 28.5 | 100 | 30.5 | 129 | 29.1 | 126 |
| F9 | BA.1.1 | Mar 2022 | 29.1 | 36 | 29.2 | 14.8 | 29.5 | 10.9 |
| F10 | BA.1 | Jan 2022 | 29.9 | 4.29 | 31.5 | 4.76 | 31.8 | 2.89 |
| F11 | BA.1 | Jan 2022 | 31 | 0.7 | 34.6 | 0.290 | 32.7 | 0.000 |
| F12 | BA.1.1 | Jan 2022 | 31.4 | 0.462 | 35.2 | 1.14 | 34.9 | 0.642 |
| F13 | BA.1 | Jan 2022 | 31.9 | 0 | 33.4 | 0 | 34.8 | 0 |
| F14 | BA.1.1 | Feb 2022 | 32.6 | 0 | 34.4 | 0 | 34.4 | 0 |

| | | | | | | | | |
|---|---|---|---|---|---|---|---|---|
| F15 | BA.1.1 | Jan 2022 | 33.8 | 0 | 37.3 | 0 | 37.1 | 0 |
| F16 | BA.1.1 | Jan 2022 | 34.5 | 0 | 34.2 | 0 | QNS | 0 |

FT1=Freeze thaw 1, FT2=Freeze thaw 2, Conc=Concentration

**Table S3**. Univariate and multivariate logistic regression analyses between key predictors and having a positive Calu-3 infectivity assay result for Delta and Omicron samples.

| **Characteristic** | **OR (95% CI)** | **p-value** | **OR (95% CI)** | **p-value** |
|---|---|---|---|---|
| **Delta, N=75** | **Univariate** | | **Multivariate (N=73)** | |
| $C_T$ | 0.75 (0.63, 0.88) | <0.001 | 0.72 (0.58, 0.86) | 0.001 |
| Antigen concentration, pg/ml | 1.00 (1.00, 1.00) | 0.95 | 1.00 (1.00, 1.00) | 0.20 |
| Symptom Duration, days | 0.85 (0.71, 1.00) | 0.06 | 0.89 (0.72, 1.09) | 0.25 |
| Missing | 2 | | | |
| Age, years | 0.99 (0.96, 1.03) | 0.72 | 1.02 (0.98, 1.06) | 0.42 |
| Missing | 1 | | | |
| Vaccine Status | | | | |
| Not fully vaccinated | Ref | | Ref | |
| At least fully vaccinated | 0.62 (0.25, 1.53) | 0.30 | 0.44 (0.13, 1.32) | 0.15 |
| **Omicron, N=85** | **Univariate** | | **Multivariate (N=85)** | |
| $C_T$ | 1.03 (0.90, 1.17) | 0.68 | 1.04 (0.88, 1.23) | 0.65 |
| Antigen concentration, pg/ml | 1.00 (1.00, 1.00) | 0.91 | 1.00 (1.00, 1.00) | 0.83 |
| Symptom Duration, days | 1.00 (0.97, 1.03) | 0.99 | 0.95 (0.72, 1.25) | 0.72 |
| Age, years | 0.97 (0.74, 1.27) | 0.82 | 1.00 (0.97, 1.04) | 0.85 |
| Vaccine Status | | | | |
| Not fully vaccinated | Ref | | Ref | |
| At least fully vaccinated | 0.80 (0.34, 1.89) | 0.61 | 0.71 (0.24, 2.04) | 0.53 |

OR = Odds Ratio, CI = Confidence Interval, Ref = Reference Level

**Table S4. Univariate and multivariate logistic regression analyses between key predictors and having a positive TMPRSS-2 infectivity assay result for Delta and Omicron samples**.

| Characteristic | OR (95% CI) | p-value | OR (95% CI) | p-value |
|---|---|---|---|---|
| **Delta** | **Univariate** | | **Multivariate (N=73)** | |
| $C_T$ | 0.70 (0.52, 0.88) | 0.007 | 0.70 (0.49, 0.92) | 0.02 |
| Antigen concentration, pg/ml | 1.00 (1.00, 1.00) | 0.52 | 1.00 (1.00, 1.00) | 0.63 |
| Symptom Duration, days | 0.79 (0.64, 0.94) | 0.01 | 0.82 (0.63, 1.05) | 0.11 |
| Missing | 2 | | | |
| Age, years | 0.97 (0.92, 1.01) | 0.13 | 1.00 (0.94, 1.06) | 0.88 |
| Missing | 1 | | | |
| Vaccine Status | | | | |
| Not fully vaccinated | Ref | | Ref | |
| At least fully vaccinated | 0.77 (0.23, 2.49) | 0.67 | 0.40 (0.08, 1.76) | 0.25 |
| **Omicron** | **Univariate** | | **Multivariate (N=85)** | |
| $C_T$ | 1.11 (0.98, 1.28) | 0.12 | 1.11 (0.94, 1.33) | 0.24 |
| Antigen concentration, pg/ml | 1.00 (1.00, 1.00) | 0.36 | 1.00 (1.00, 1.00) | 0.99 |
| Symptom Duration, days | 1.02 (0.77, 1.34) | 0.89 | 0.95 (0.71, 1.27) | 0.74 |
| Age, years | 1.01 (0.98, 1.04) | 0.66 | 1.02 (0.98, 1.06) | 0.40 |
| Vaccine Status | | | | |
| Not fully vaccinated | Ref | | Ref | |
| At least fully vaccinated | 0.54 (0.22, 1.30) | 0.17 | 0.35 (0.11, 1.08) | 0.08 |

OR = Odds Ratio, CI = Confidence Interval, Ref = Reference Level

**Table S5. Twenty archived positive Omicron & Delta samples were selected and tested in parallel on both XPRSARS-COV2-10 and XP3COV2/FLU/RSV-10 cartridges to verify that the E target was the appropriate comparator.**

| ID | Lineage | Original Assay Test Date | Date Study Performed | XPRSARS-COV2-10 (E Target $C_T$) | XPRCOV2/FLU/RSV-10 (COVID Target $C_T$) | Target $C_T$ Difference (COVID $C_T$ minus E $C_T$) |
|---|---|---|---|---|---|---|
| B1 | Unknown | Jan 2022 | June 2022 | 0 (N2 42.3) | 0 | 0 |
| B2 | BA.1.1 | Jan 2022 | June 2022 | 31.2 | 31.5 | 0.3 |
| B3 | BA.1.1 | Jan 2022 | June 2022 | 31.6 | 32.3 | 0.7 |
| B4 | BA.1 | Jan 2022 | June 2022 | 28.1 | 30.7 | 2.6 |
| B5 | BA.1 | Dec 2021 | June 2022 | 20.3 | 20.4 | 0.1 |
| B6 | BA.1 | Jan 2022 | June 2022 | 23.8 | 23.9 | 0.1 |
| B7 | BA.1 | Jan 2022 | June 2022 | 24.5 | 24.8 | 0.3 |
| B8 | AY.25 | Jan 2022 | June 2022 | 25.2 | 25.9 | 0.7 |
| B9 | BA.1.1 | Jan 2022 | June 2022 | 32.1 | 32.8 | 0.7 |
| B10 | BA.1 | Jan 2022 | June 2022 | 39 | 40 | 1 |
| B11 | BA.1 | Jan 2022 | June 2022 | 32.7 | 33.5 | 0.8 |
| B12 | BA.1.1 | Jan 2022 | June 2022 | 30.4 | 30.7 | 0.3 |
| B13 | BA.1.1 | Jan 2022 | June 2022 | 28.2 | 30.5 | 2.3 |
| B14 | BA.1 | Jan 2022 | June 2022 | 31.4 | 35.1 | 3.7 |

| | | | | | | |
|---|---|---|---|---|---|---|
| B15 | BA.1 | Jan 2022 | June 2022 | 28.3 | 27.6 | -0.7 |
| B16 | BA.1.1 | Jan 2022 | June 2022 | 25.5 | 26.8 | 1.3 |
| B17 | BA.1.1 | Jan 2022 | June 2022 | 36.2 | 35.5 | -0.7 |
| B18 | BA.1 | Jan 2022 | June 2022 | 40.2 | 36.1 | -4.1 |
| B19 | BA.1.1 | Jan 2022 | June 2022 | 30.9 | 30.4 | -0.5 |
| B20 | BA.1.1 | Jan 2022 | June 2022 | 24.5 | 25 | 0.5 |

**Supplementary Data File: Remnant clinical sample information**. For the 334 remnant clinical samples used in this study, this data file reports: demographic and clinical information including vaccine status and symptoms; results from antigen and PCR testing; results from rapid antigen testing; sequencing results; and results from infectivity assays.
